# COVID-19-associated hospitalizations among vaccinated and unvaccinated adults ≥18 years – COVID-NET, 13 states, January 1 – July 24, 2021

**DOI:** 10.1101/2021.08.27.21262356

**Authors:** Fiona P. Havers, Huong Pham, Christopher A. Taylor, Michael Whitaker, Kadam Patel, Onika Anglin, Anita K. Kambhampati, Jennifer Milucky, Elizabeth Zell, Shua J. Chai, Pam Daily Kirley, Nisha B. Alden, Isaac Armistead, Kimberly Yousey-Hindes, James Meek, Kyle P. Openo, Evan J. Anderson, Libby Reeg, Alexander Kohrman, Ruth Lynfield, Kathryn Como-Sabetti, Elizabeth M. Davis, Cory Cline, Alison Muse, Grant Barney, Sophrena Bushey, Christina B. Felsen, Laurie M. Billing, Eli Shiltz, Melissa Sutton, Nasreen Abdullah, H. Keipp Talbot, William Schaffner, Mary Hill, Andrea George, Bhavini Patel Murthy, Meredith McMorrow

## Abstract

**Background:** As of August 21, 2021, >60% of the U.S. population aged ≥18 years were fully vaccinated with vaccines highly effective in preventing hospitalization due to Coronavirus Disease-2019 (COVID-19). Infection despite full vaccination (vaccine breakthrough) has been reported, but characteristics of those with vaccine breakthrough resulting in hospitalization and relative rates of hospitalization in unvaccinated and vaccinated persons are not well described, including during late June and July 2021 when the highly transmissible Delta variant predominated.

**Methods:** From January 1–June 30, 2021, cases defined as adults aged ≥18 years with laboratory-confirmed Severe Acute Respiratory Coronavirus-2 (SARS-CoV-2) infection were identified from >250 acute care hospitals in the population-based COVID-19-Associated Hospitalization Surveillance Network (COVID-NET). Through chart review for sampled cases, we examine characteristics associated with vaccination breakthrough. From January 24–July 24, 2021, state immunization information system data linked to both >37,000 cases representative cases and the defined surveillance catchment area population were used to compare weekly hospitalization rates in vaccinated and unvaccinated individuals. Unweighted case counts and weighted percentages are presented.

**Results:** From January 1 – June 30, 2021, fully vaccinated cases increased from 1 (0.01%) to 321 (16.1%) per month. Among 4,732 sampled cases, fully vaccinated persons admitted with COVID-19 were older compared with unvaccinated persons (median age 73 years [Interquartile Range (IQR) 65-80] v. 59 years [IQR 48-70]; p<0.001), more likely to have 3 or more underlying medical conditions (201 (70.8%) v. 2,305 (56.1%), respectively; p<0.001) and be residents of long-term care facilities [37 (14.5%) v. 146 (5.5%), respectively; p<0.001]. From January 24 – July 24, 2021, cumulative hospitalization rates were 17 times higher in unvaccinated persons compared with vaccinated persons (423 cases per 100,000 population v. 26 per 100,000 population, respectively); rate ratios were 23, 22 and 13 for those aged 18-49, 50-64, and ≥65 years respectively. For June 27 – July 24, hospitalization rates were ≥10 times higher in unvaccinated persons compared with vaccinated persons for all age groups across all weeks.

**Conclusion:** Population-based hospitalization rates show that unvaccinated adults aged ≥18 years are 17 times more likely to be hospitalized compared with vaccinated adults. Rates are far higher in unvaccinated persons in all adult age groups, including during a period when the Delta variant was the predominant strain of the SARS-CoV-2 virus. Vaccines continue to play a critical role in preventing serious COVID-19 illness and remain highly effective in preventing COVID-19 hospitalizations.

## Introduction

As of August 21, 2021, 170 million people in the United States were fully vaccinated against COVID-19, including >62% of the population aged ≥18 years.^2^ Consistent with other studies, data from the Centers for Disease Control and Prevention’s (CDC) Coronavirus Disease 2019-Associated Hospitalization Surveillance Network (COVID-NET) have previously shown that all three vaccine products currently available are highly effective in preventing hospitalization in adults aged ≥65 years.^3,4^ Infections in fully vaccinated persons (i.e., vaccine breakthroughs cases) are expected,^5^ even in the setting of highly effective vaccines. While most infections occurring in fully vaccinated individuals have been mild or asymptomatic,^5^ more serious SARS-CoV-2 infections can occur in fully vaccinated individuals.^4^ However, the characteristics of fully vaccinated patients hospitalized with COVID-19 and factors associated with risk of vaccine breakthrough have not been well described. Using COVID-NET data representing >67,000 COVID-19-associated hospitalizations from January–June 2021, we compare characteristics of persons hospitalized with laboratory-confirmed SARS-CoV-2 virus infection by vaccination status and assess characteristics of hospitalized cases with vaccination breakthrough. We also compare clinical outcomes between unvaccinated and fully vaccinated persons admitted for COVID-19-associated illness. We use state immunization information system (IIS) linked to both hospitalized cases with laboratory-confirmed SARS-CoV-2 infection and the population of the defined COVID-NET catchment area to compare weekly population-based hospitalization rates in vaccinated and unvaccinated individuals, including for late June and July 2021, a period when the highly-transmissible Delta variant was the predominant strain circulating.^6^

## Methods

### COVID-NET description and data collected for all COVID-NET Cases

COVID-NET is a population-based surveillance system of laboratory-confirmed COVID-19-associated hospitalizations comprising 99 counties in 14 states (California, Colorado, Connecticut, Georgia, Iowa, Maryland, Michigan, Minnesota, New Mexico, New York, Ohio, Oregon, Tennessee, and Utah) and covering approximately 10% of the US population. Hospitalized patients residing in a surveillance catchment area with a positive molecular or rapid antigen detection test for SARS-CoV-2 during hospitalization or within 14 days before admission are included as COVID-NET cases.^7^ One site (Iowa) does not have access to the state IISs and was excluded from this analysis.

Basic demographic information, including age, race and Hispanic ethnicity, sex, hospital admission date, and evidence of a positive SARS-CoV-2 test are transmitted weekly on all patients.^7^ Since few children were fully vaccinated during the analysis period, this analysis was limited to adult cases aged ≥18 years.

### COVID-NET Sampling/Weighting Methodology

Detailed data on patient demographics, COVID-19 vaccination status, primary reason for admission, select underlying medical conditions, and clinical outcomes, including ICU admission and death, are collected on a representative sample of patients stratified by age and site. To select sampled cases, random numbers are generated and assigned to each case. Sampling weights are based on the probability of selection and adjusted for non-response; sample sizes vary by surveillance month, site and age group and are based on the total number of cases identified in each of these strata.^8^ Monthly demographic and clinical data through June 2021 were the most recent data available at the time of this study.

### Vaccination definitions, weighting of cases with known vaccination status

Fully vaccinated cases were defined as having a positive SARS-CoV-2 test ≥14 days after either the second dose of a 2-dose series or after one dose of a single dose series. Partial vaccination was defined as ≥14 days after a single dose through <14 days after the second dose in a 2-dose series. If the SARS-CoV-2 test date was not available, hospital admission date was used. Partially vaccinated patients and patients with positive tests <14 days after one vaccine dose, regardless of type, were excluded from further analyses.

Vaccination status for both hospitalized cases and the underlying population was determined by state IIS data, as previously described.^3^ We collected COVID-19 vaccination status (doses, dates administered, and product) from state IISs for all sampled COVID-NET cases. Some sites expanded collection of vaccination status to non-sampled cases, which were included for analysis of demographic factors and population-based rates associated with vaccination if all cases in a single month and age group had vaccination status available (“vaccine sample”).

For fully vaccinated sampled cases only, COVID-NET personnel reviewed state health department electronic laboratory records and other data sources for a positive SARS-CoV-2 tests predating the test associated with the current hospitalization. This was done to exclude those who were reinfected or had prolonged viral shedding from a prior SARS-CoV-2 infection that predated full vaccination and, therefore, did not meet the definition of vaccine breakthrough **(Supplementary Table 2)**. Data on prior positive SARS-CoV-2 tests were not available for unvaccinated persons.

### Comparison of Clinical Characteristics and Outcomes among a Weighted Sample of Fully Vaccinated and Unvaccinated Hospitalized Cases with COVID-19

For all sampled cases, trained surveillance staff conduct medical chart abstractions using a standard case report form. We compared demographic information, underlying medical conditions, clinical outcomes, signs and symptoms at admission, and reasons for admission between unvaccinated and fully vaccinated sampled cases (“comparison sample”). Underlying medical conditions were categorized into major groups **(Supplementary Table 1)**. Two physicians reviewed reason for admission and symptoms, and those cases whose reason for admission might not have been primarily related to COVID-19, despite receiving a positive SARS-CoV-2 laboratory test, were excluded from the comparison sample.^3^

We used multivariable logistic regression to examine factors associated with vaccine breakthrough cases; models included age, race and Hispanic ethnicity, long-term care facility (LTCF) residence, and underlying medical conditions. We also used multivariable logistic regression to examine the association of vaccination with severe COVID-19, defined as ICU admission or in-hospital death. Because older adults were eligible for vaccination earlier than other age groups^9^ and older age has been associated with increased risk of vaccine breakthrough with other infections, we stratified all models, analyzing those aged <65 and ≥65 years separately.

### Description of Population-Based COVID-19-Associated Hospitalization Rates among Unvaccinated and Vaccinated Persons

We collected a minimum set of data on all identified cases to produce weekly hospitalization rates (https://gis.cdc.gov/grasp/COVIDNet/COVID19_3.html) for all COVID-NET cases. Incidence rates were calculated using the National Center for Health Statistics’ vintage 2019 bridge-race postcensal population estimates for the counties included in COVID-NET surveillance.^10^ To determine population-based rates of hospitalization by vaccination status per 100,000 persons ≥18 years of age, county-level coverage in the COVID-NET catchment area was estimated using population denominators. Vaccination status was classified as described above for hospitalized cases using the vaccine sample; weekly population-based rates were calculated based on denominators that changed weekly depending on the vaccination status of individuals in the underlying population. Rate ratios were calculated for all fully vaccinated and unvaccinated cases who met the case definition, regardless of prior SARS-CoV-2 test results or reason for admission; overall rates for those ≥18 years were standardized to the underlying population. We excluded partially vaccinated persons from rate calculations. For analyses comparing hospitalization rates in unvaccinated with vaccinated persons only, data through the week ending July 24, 2021 were included, as they were the most recent data available at the time of this study. Weeks in which the Delta variant was estimated to cause >50% of new SARS-COV-2 infections, June 27 – July 24, 2021,^6^ were examined. A method for formally calculating vaccine effectiveness from COVID-NET data has been published;^11^ VE estimates will be included in a forthcoming manuscript.

### Statistical Analysis

Data from all cases aged ≥18 years and hospitalized with laboratory-confirmed COVID-19 during January 1–June 30, 2021 with linked vaccine registry data were used to describe the vaccination status of hospitalized cases by age, sex, race and Hispanic ethnicity, and month of admission. The comparison sample used in multivariable models included age groups (18–49, 50–64, ≥65 years), sex, and race and ethnicity; models incorporated clustering by site to account for geographic differences. Other variables with p-values <0.10 in bivariate analyses were included in the multivariable analyses. Model fit was assessed with Quasi-likelihood under Independence Model Criterion (QIC). Log-linked Poisson generalized estimating equations regression with an independent working correlation structure was used to generate adjusted risk ratios (aRR) and 95% confidence intervals (CI). Data were analyzed using SAS survey procedures to account for sampling weights. Unweighted case counts and weighted percentages are presented unless otherwise noted. Proportions by month with 95% confidence intervals are presented for binary measures, and medians with interquartile ranges for continuous measures. We used Taylor series linearization methods for variance estimation. All analyses were conducted using SAS 9.4 software (SAS Institute Inc., Cary, NC).

This activity was reviewed by CDC and was conducted consistent with applicable federal law and CDC policy (see e.g., 45 C.F.R. part 46.102(l)(2), 21 C.F.R. part 56; 42 U.S.C. §241(d); 5 U.S.C.

## Results

During January 1–June 30, 2021, there were 67,311 laboratory-confirmed COVID-19-associated hospitalized cases ≥18 years identified in COVID-NET and meeting the case definition, among whom a representative sample of 35,846 had vaccination data linked to state vaccination registries (vaccine sample) **(Supplementary Figure 1)**. Among these, 30,967 (87.6% (weighted percentages shown throughout)) were unvaccinated, 1,791 (5.0%) had received 1 dose <14 d prior to admission, 1,753 (4.4%) were partially vaccinated and 1,255 (3%) were fully vaccinated (**Table 1)**. The number of cases that were fully vaccinated increased from 1 (0.0%) in January to 321 (16.1%) in June. A greater proportion of cases in older age groups were fully vaccinated; 32% of hospitalized cases aged ≥65 years in June were fully vaccinated **(Supplementary Figure 2)**.

**Table 1.**
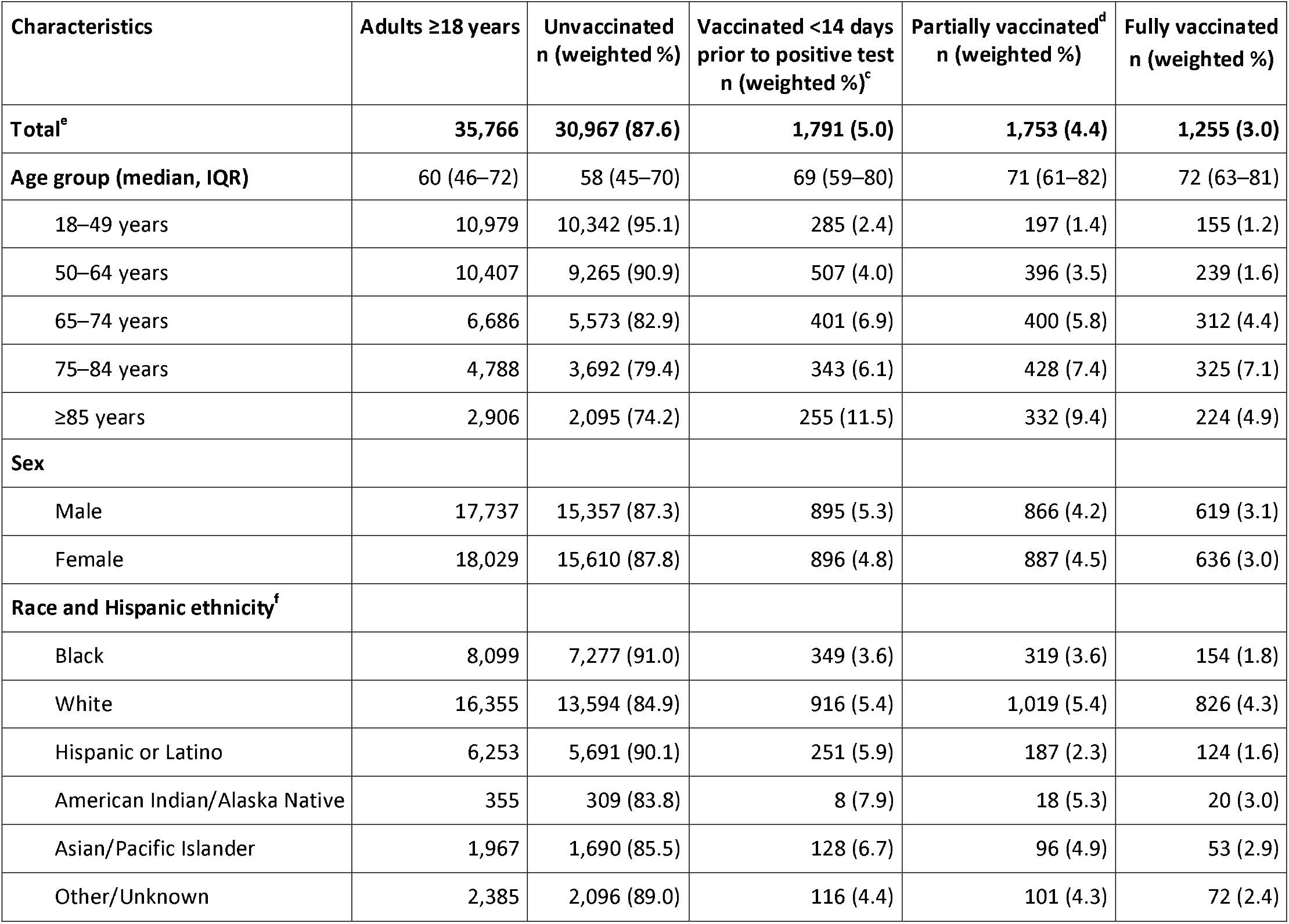
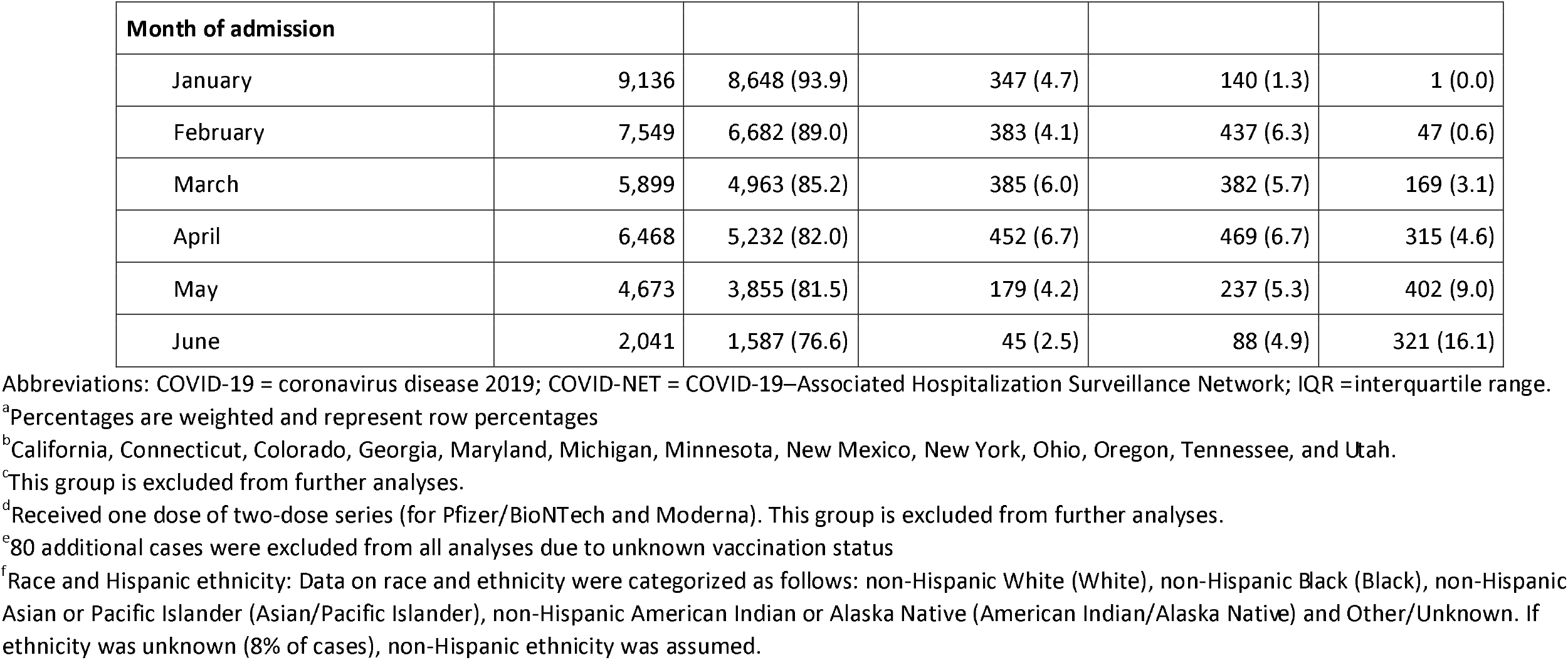
Demographics of a representative sample^a^ of hospitalized adults aged ≥18 years with laboratory-confirmed COVID-19-associated hospitalization admitted January 1–June 30, 2021, by vaccination status — COVID-NET, 13 States^b^.

Among a representative sample of 6,115 hospitalized adults aged ≥18 years, the following were excluded (unweighted percentages shown for excluded cases): 282 (4.6%) who had received one dose of vaccine <14 days prior to the positive SARS-CoV-2 test, 286 (4.7%) who were partially vaccinated, 14 (0.2%) with unknown vaccination status, and 33 (0.5%) cases with incomplete data. Among remaining the cases (n=5,550), 390 were fully vaccinated and 5,160 were unvaccinated; among fully vaccinated cases, 52/390 (13%) had evidence of prior SARS-CoV-2 positive tests and were also excluded. An additional 716 (13%) of the remaining 5,448 cases were excluded as their reason for admission might not have been primarily related to COVID-19; the proportion of cases excluded for this reason was similar among the remaining vaccinated cases compared with unvaccinated cases ((46/338 (14%) v. 670/5110 (13%), respectively; p-value = 0.79)). **(Supplementary Figure 1)**.

The comparison sample was thus restricted to the remaining 4,732 cases with hospitalization associated primarily with COVID-19 illness, including 4,440 unvaccinated and 292 fully vaccinated cases **(Table 2)**. Among fully vaccinated cases, the median time from full vaccination status until hospital admission was 42 days (IQR 28–68). Among all adults aged ≥18 years, fully vaccinated cases were older when compared to unvaccinated cases [median age 73 years (IQR 65-80) v. 59 years (IQR 48-70); p<0.001] and more likely to be LTCF residents [37 (14.5%) v. 146 (5.5%), respectively; p<0.001]. In addition, 69 (32.1%) fully vaccinated cases were more likely to be immunosuppressed compared with 442 (11.3%) unvaccinated cases (p<0.001); these fully vaccinated immunosuppressed cases included 25 (36%) and 14 (20%) cases with solid organ malignancy and a history of a solid organ transplant, respectively. Vaccinated cases were also more likely to have 3 or more underlying medical conditions compared with unvaccinated cases (201 (70.8%) v. 2,305 (56.1%), respectively; p<0.001).

**Table 2.**
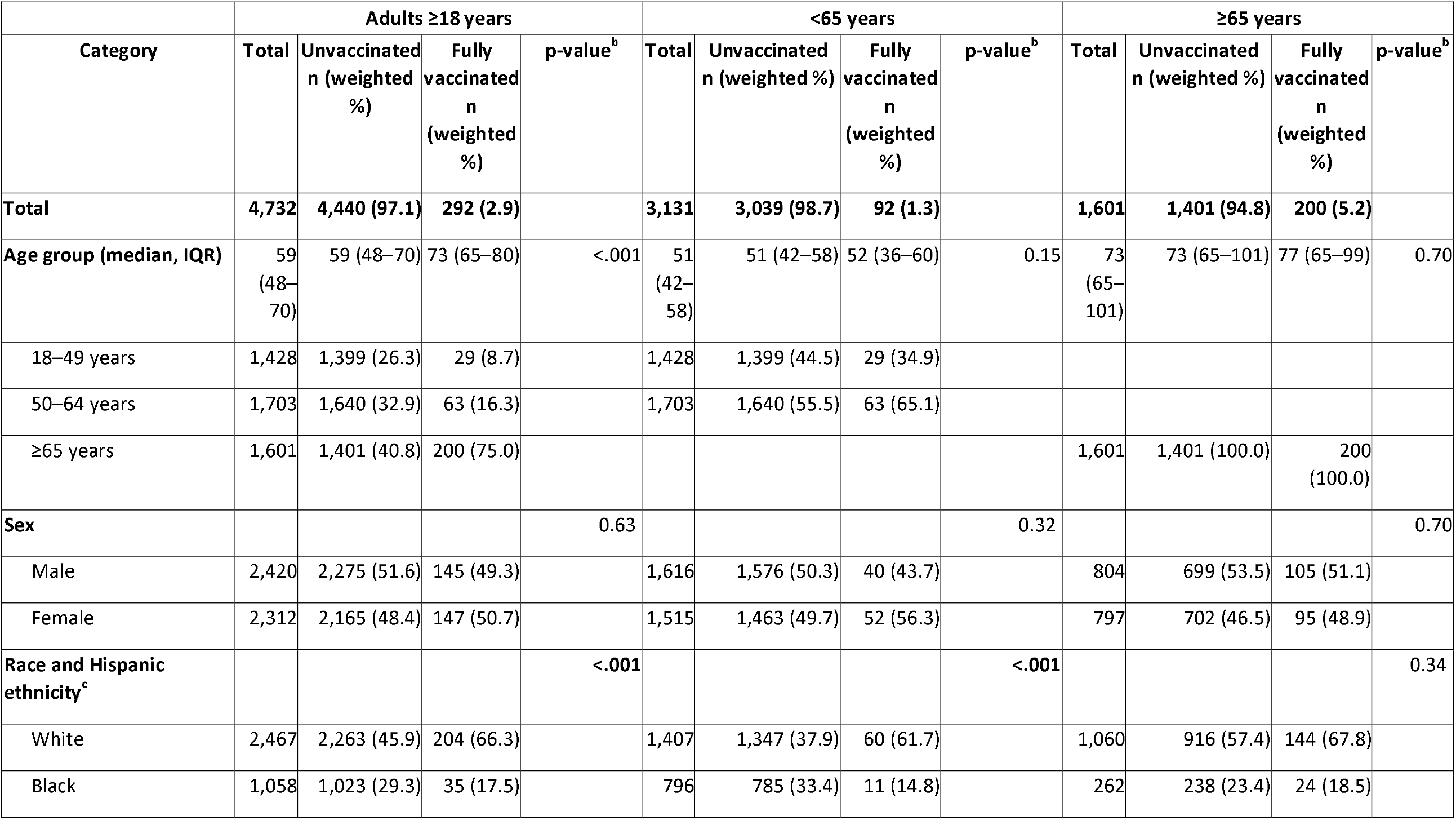
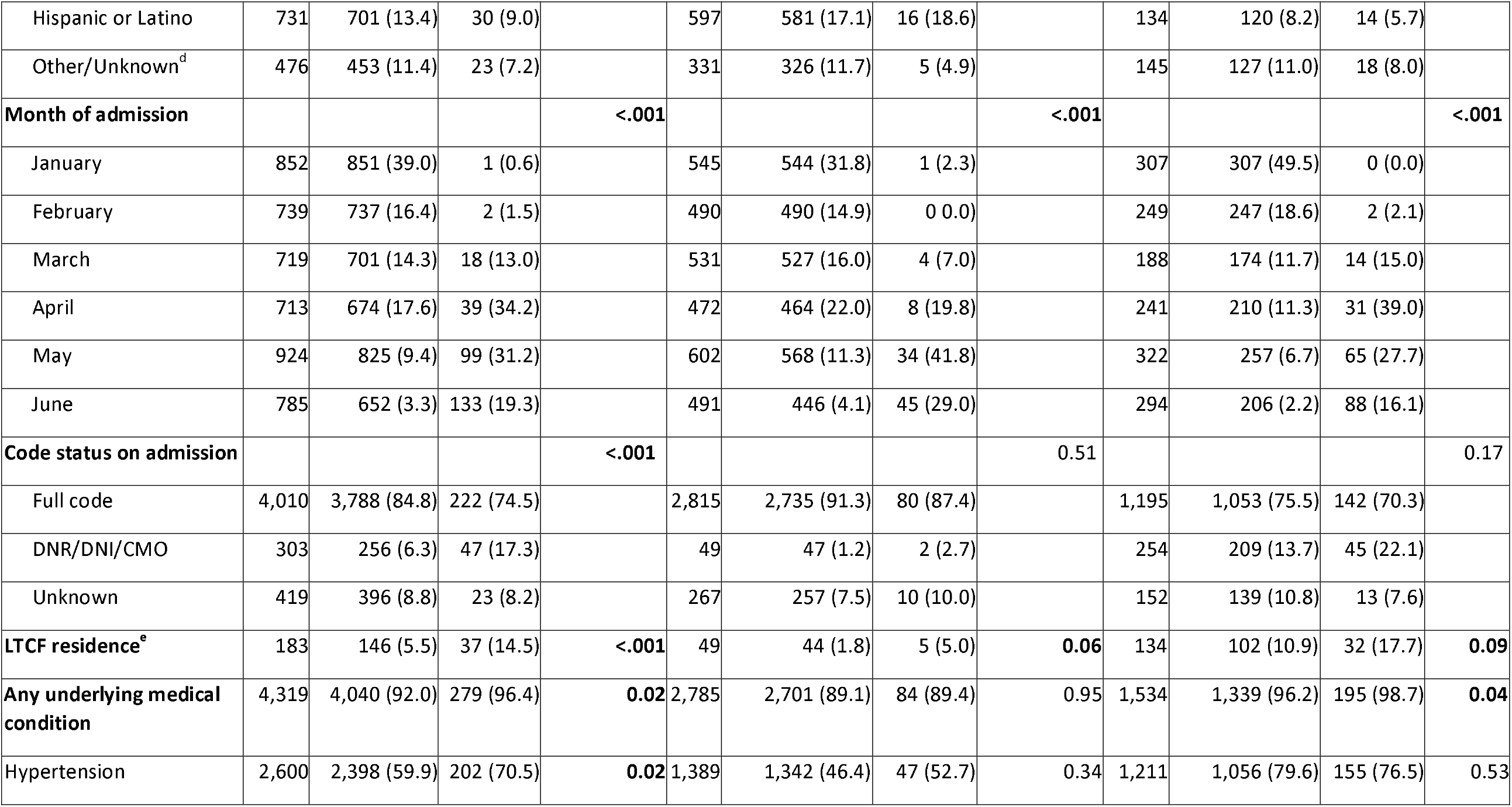
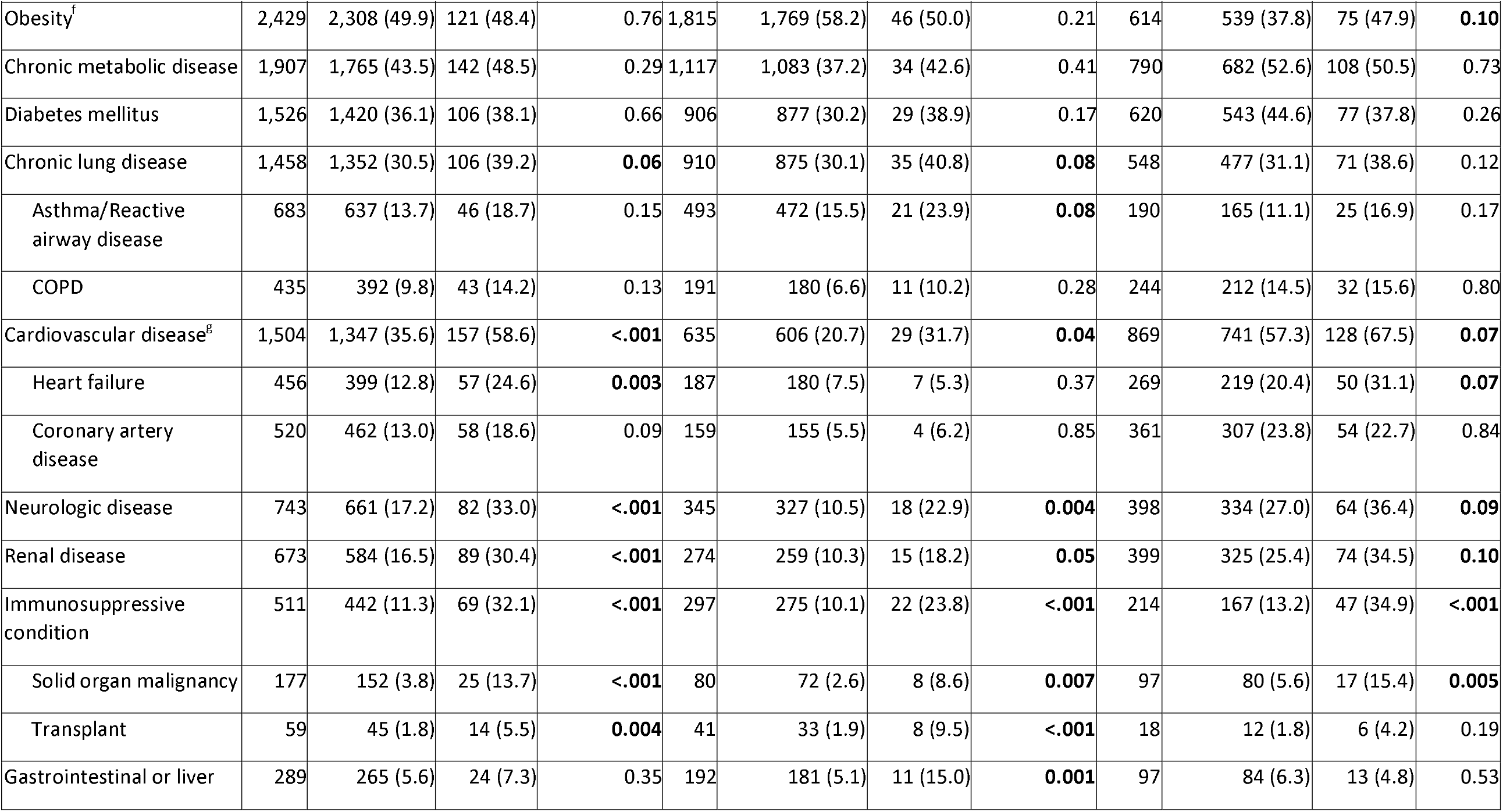
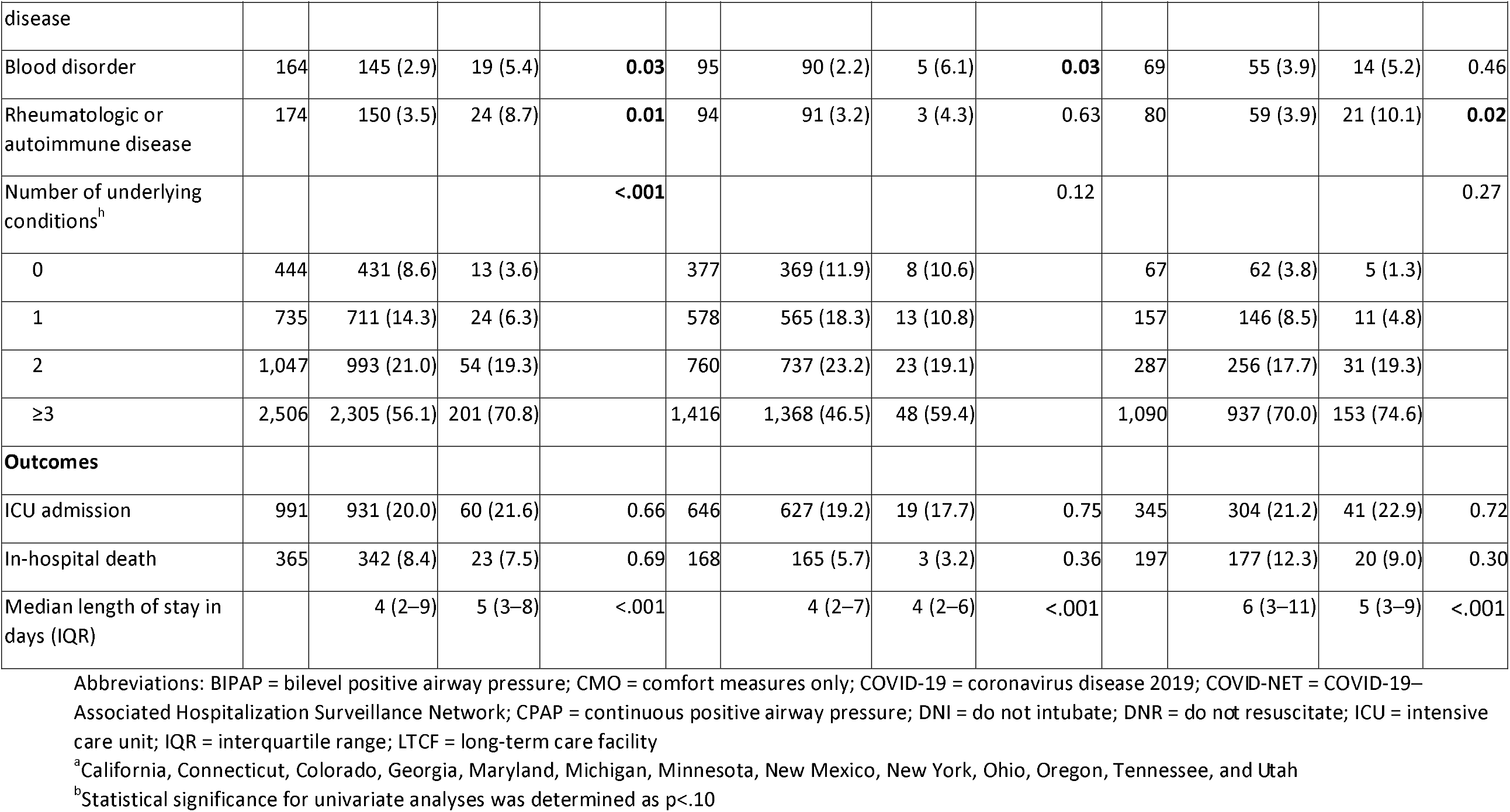
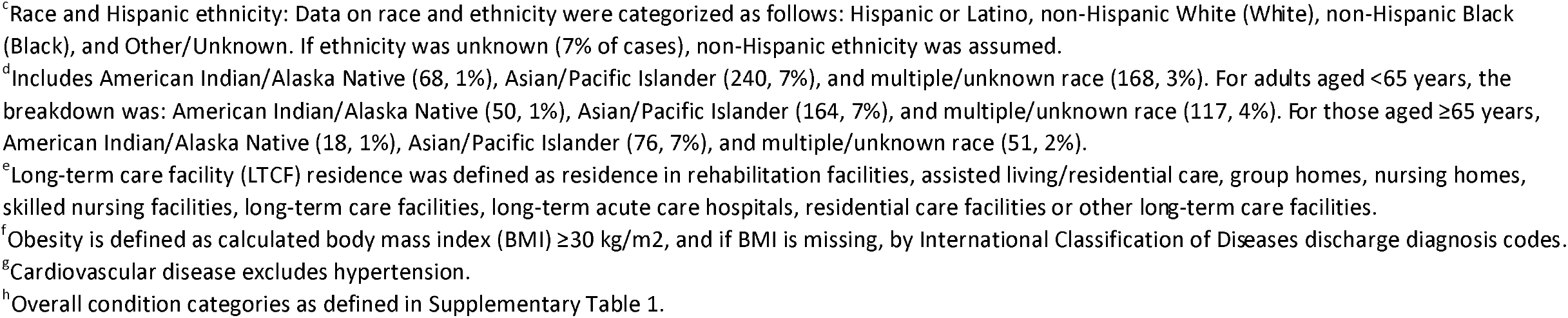
Demographics, underlying conditions, reason for admission, and clinical outcomes among a representative sample of unvaccinated and fully vaccinated hospitalized adults aged ≥18 years with laboratory-confirmed COVID-19-associated hospitalization admitted January 1– June 30, 2021, stratified by age (<65 and ≥65 years) and restricted to those with COVID-19-associated hospitalization as the likely primary reason for admission — COVID-NET, 13 States^a^.

| Category | Adults ≥18 years |  |  |  | <65 years |  |  |  | ≥65 years |  |  |  |
| --- | --- | --- | --- | --- | --- | --- | --- | --- | --- | --- | --- | --- |
|  | Total | Unvaccinated<br>n (weighted<br>%) | Fully<br>vaccinated<br>n<br>(weighted<br>%) | p-value<sup>b</sup> | Total | Unvaccinated<br>n (weighted %) | Fully<br>vaccinated<br>n<br>(weighted<br>%) | p-value<sup>b</sup> | Total | Unvaccinated<br>n (weighted %) | Fully<br>vaccinated<br>n<br>(weighted<br>%) | p-value<sup>b</sup> |
| **Total** | **4,732** | **4,440 (97.1)** | **292 (2.9)** |  | **3,131** | **3,039 (98.7)** | **92 (1.3)** |  | **1,601** | **1,401 (94.8)** | **200 (5.2)** |  |
| **Age group (median, IQR)** | 59<br>(48–70) | 59 (48–70) | 73 (65–80) | <.001 | 51<br>(42–58) | 51 (42–58) | 52 (36–60) | 0.15 | 73<br>(65–101) | 73 (65–101) | 77 (65–99) | 0.70 |
| 18–49 years | 1,428 | 1,399 (26.3) | 29 (8.7) |  | 1,428 | 1,399 (44.5) | 29 (34.9) |  |  |  |  |  |
| 50–64 years | 1,703 | 1,640 (32.9) | 63 (16.3) |  | 1,703 | 1,640 (55.5) | 63 (65.1) |  |  |  |  |  |
| ≥65 years | 1,601 | 1,401 (40.8) | 200 (75.0) |  |  |  |  |  | 1,601 | 1,401 (100.0) | 200 (100.0) |  |
| **Sex** |  |  |  | 0.63 |  |  |  | 0.32 |  |  |  | 0.70 |
| Male | 2,420 | 2,275 (51.6) | 145 (49.3) |  | 1,616 | 1,576 (50.3) | 40 (43.7) |  | 804 | 699 (53.5) | 105 (51.1) |  |
| Female | 2,312 | 2,165 (48.4) | 147 (50.7) |  | 1,515 | 1,463 (49.7) | 52 (56.3) |  | 797 | 702 (46.5) | 95 (48.9) |  |
| **Race and Hispanic ethnicity<sup>c</sup>** |  |  |  | <.001 |  |  |  | <.001 |  |  |  | 0.34 |
| White | 2,467 | 2,263 (45.9) | 204 (66.3) |  | 1,407 | 1,347 (37.9) | 60 (61.7) |  | 1,060 | 916 (57.4) | 144 (67.8) |  |
| Black | 1,058 | 1,023 (29.3) | 35 (17.5) |  | 796 | 785 (33.4) | 11 (14.8) |  | 262 | 238 (23.4) | 24 (18.5) |  |
|  | Total | Unvaccinated<br>n (weighted<br>%) | Fully<br>vaccinated<br>n<br>(weighted<br>%) | p-value<sup>b</sup> | Total | Unvaccinated<br>n (weighted %) | Fully<br>vaccinated<br>n<br>(weighted<br>%) | p-value<sup>b</sup> | Total | Unvaccinated<br>n (weighted %) | Fully<br>vaccinated<br>n<br>(weighted<br>%) | p-value<sup>b</sup> |
| Hispanic or Latino | 731 | 701 (13.4) | 30 (9.0) |  | 597 | 581 (17.1) | 16 (18.6) |  | 134 | 120 (8.2) | 14 (5.7) |  |
| Other/Unknown<sup>d</sup> | 476 | 453 (11.4) | 23 (7.2) |  | 331 | 326 (11.7) | 5 (4.9) |  | 145 | 127 (11.0) | 18 (8.0) |  |
| **Month of admission** |  |  |  | **<.001** |  |  |  | **<.001** |  |  |  | **<.001** |
| January | 852 | 851 (39.0) | 1 (0.6) |  | 545 | 544 (31.8) | 1 (2.3) |  | 307 | 307 (49.5) | 0 (0.0) |  |
| February | 739 | 737 (16.4) | 2 (1.5) |  | 490 | 490 (14.9) | 0 (0.0) |  | 249 | 247 (18.6) | 2 (2.1) |  |
| March | 719 | 701 (14.3) | 18 (13.0) |  | 531 | 527 (16.0) | 4 (7.0) |  | 188 | 174 (11.7) | 14 (15.0) |  |
| April | 713 | 674 (17.6) | 39 (34.2) |  | 472 | 464 (22.0) | 8 (19.8) |  | 241 | 210 (11.3) | 31 (39.0) |  |
| May | 924 | 825 (9.4) | 99 (31.2) |  | 602 | 568 (11.3) | 34 (41.8) |  | 322 | 257 (6.7) | 65 (27.7) |  |
| June | 785 | 652 (3.3) | 133 (19.3) |  | 491 | 446 (4.1) | 45 (29.0) |  | 294 | 206 (2.2) | 88 (16.1) |  |
| **Code status on admission** |  |  |  | **<.001** |  |  |  | 0.51 |  |  |  | 0.17 |
| Full code | 4,010 | 3,788 (84.8) | 222 (74.5) |  | 2,815 | 2,735 (91.3) | 80 (87.4) |  | 1,195 | 1,053 (75.5) | 142 (70.3) |  |
| DNR/DNI/CMO | 303 | 256 (6.3) | 47 (17.3) |  | 49 | 47 (1.2) | 2 (2.7) |  | 254 | 209 (13.7) | 45 (22.1) |  |
| Unknown | 419 | 396 (8.8) | 23 (8.2) |  | 267 | 257 (7.5) | 10 (10.0) |  | 152 | 139 (10.8) | 13 (7.6) |  |
| **LTCF residence<sup>e</sup>** | 183 | 146 (5.5) | 37 (14.5) | **<.001** | 49 | 44 (1.8) | 5 (5.0) | **0.06** | 134 | 102 (10.9) | 32 (17.7) | **0.09** |
| **Any underlying medical condition** | 4,319 | 4,040 (92.0) | 279 (96.4) | **0.02** | 2,785 | 2,701 (89.1) | 84 (89.4) | 0.95 | 1,534 | 1,339 (96.2) | 195 (98.7) | **0.04** |
| Hypertension | 2,600 | 2,398 (59.9) | 202 (70.5) | **0.02** | 1,389 | 1,342 (46.4) | 47 (52.7) | 0.34 | 1,211 | 1,056 (79.6) | 155 (76.5) | 0.53 |
|  | Total | Unvaccinated<br>n (weighted<br>%) | Fully<br>vaccinated<br>n<br>(weighted<br>%) | p-value<sup>b</sup> | Total | Unvaccinated<br>n (weighted %) | Fully<br>vaccinated<br>n<br>(weighted<br>%) | p-value<sup>b</sup> | Total | Unvaccinated<br>n (weighted %) | Fully<br>vaccinated<br>n<br>(weighted<br>%) | p-value<sup>b</sup> |
| Obesity<sup>f</sup> | 2,429 | 2,308 (49.9) | 121 (48.4) | 0.76 | 1,815 | 1,769 (58.2) | 46 (50.0) | 0.21 | 614 | 539 (37.8) | 75 (47.9) | **0.10** |
| Chronic metabolic disease | 1,907 | 1,765 (43.5) | 142 (48.5) | 0.29 | 1,117 | 1,083 (37.2) | 34 (42.6) | 0.41 | 790 | 682 (52.6) | 108 (50.5) | 0.73 |
| Diabetes mellitus | 1,526 | 1,420 (36.1) | 106 (38.1) | 0.66 | 906 | 877 (30.2) | 29 (38.9) | 0.17 | 620 | 543 (44.6) | 77 (37.8) | 0.26 |
| Chronic lung disease | 1,458 | 1,352 (30.5) | 106 (39.2) | **0.06** | 910 | 875 (30.1) | 35 (40.8) | **0.08** | 548 | 477 (31.1) | 71 (38.6) | 0.12 |
| Asthma/Reactive<br>airway disease | 683 | 637 (13.7) | 46 (18.7) | 0.15 | 493 | 472 (15.5) | 21 (23.9) | **0.08** | 190 | 165 (11.1) | 25 (16.9) | 0.17 |
| COPD | 435 | 392 (9.8) | 43 (14.2) | 0.13 | 191 | 180 (6.6) | 11 (10.2) | 0.28 | 244 | 212 (14.5) | 32 (15.6) | 0.80 |
| Cardiovascular disease<sup>g</sup> | 1,504 | 1,347 (35.6) | 157 (58.6) | **<.001** | 635 | 606 (20.7) | 29 (31.7) | **0.04** | 869 | 741 (57.3) | 128 (67.5) | **0.07** |
| Heart failure | 456 | 399 (12.8) | 57 (24.6) | **0.003** | 187 | 180 (7.5) | 7 (5.3) | 0.37 | 269 | 219 (20.4) | 50 (31.1) | **0.07** |
| Coronary artery<br>disease | 520 | 462 (13.0) | 58 (18.6) | 0.09 | 159 | 155 (5.5) | 4 (6.2) | 0.85 | 361 | 307 (23.8) | 54 (22.7) | 0.84 |
| Neurologic disease | 743 | 661 (17.2) | 82 (33.0) | **<.001** | 345 | 327 (10.5) | 18 (22.9) | **0.004** | 398 | 334 (27.0) | 64 (36.4) | **0.09** |
| Renal disease | 673 | 584 (16.5) | 89 (30.4) | **<.001** | 274 | 259 (10.3) | 15 (18.2) | **0.05** | 399 | 325 (25.4) | 74 (34.5) | **0.10** |
| Immunosuppressive<br>condition | 511 | 442 (11.3) | 69 (32.1) | **<.001** | 297 | 275 (10.1) | 22 (23.8) | **<.001** | 214 | 167 (13.2) | 47 (34.9) | **<.001** |
| Solid organ malignancy | 177 | 152 (3.8) | 25 (13.7) | **<.001** | 80 | 72 (2.6) | 8 (8.6) | **0.007** | 97 | 80 (5.6) | 17 (15.4) | **0.005** |
| Transplant | 59 | 45 (1.8) | 14 (5.5) | **0.004** | 41 | 33 (1.9) | 8 (9.5) | **<.001** | 18 | 12 (1.8) | 6 (4.2) | 0.19 |
| Gastrointestinal or liver | 289 | 265 (5.6) | 24 (7.3) | 0.35 | 192 | 181 (5.1) | 11 (15.0) | **0.001** | 97 | 84 (6.3) | 13 (4.8) | 0.53 |
|  | Total | Unvaccinated<br>n (weighted<br>%) | Fully<br>vaccinated<br>n<br>(weighted<br>%) | p-value<sup>b</sup> | Total | Unvaccinated<br>n (weighted %) | Fully<br>vaccinated<br>n<br>(weighted<br>%) | p-value<sup>b</sup> | Total | Unvaccinated<br>n (weighted %) | Fully<br>vaccinated<br>n<br>(weighted<br>%) | p-value<sup>b</sup> |
| disease |  |  |  |  |  |  |  |  |  |  |  |  |
| Blood disorder | 164 | 145 (2.9) | 19 (5.4) | **0.03** | 95 | 90 (2.2) | 5 (6.1) | **0.03** | 69 | 55 (3.9) | 14 (5.2) | 0.46 |
| Rheumatologic or<br>autoimmune disease | 174 | 150 (3.5) | 24 (8.7) | **0.01** | 94 | 91 (3.2) | 3 (4.3) | 0.63 | 80 | 59 (3.9) | 21 (10.1) | **0.02** |
| Number of underlying<br>conditions<sup>h</sup> |  |  |  | **<.001** |  |  |  | 0.12 |  |  |  | 0.27 |
| 0 | 444 | 431 (8.6) | 13 (3.6) |  | 377 | 369 (11.9) | 8 (10.6) |  | 67 | 62 (3.8) | 5 (1.3) |  |
| 1 | 735 | 711 (14.3) | 24 (6.3) |  | 578 | 565 (18.3) | 13 (10.8) |  | 157 | 146 (8.5) | 11 (4.8) |  |
| 2 | 1,047 | 993 (21.0) | 54 (19.3) |  | 760 | 737 (23.2) | 23 (19.1) |  | 287 | 256 (17.7) | 31 (19.3) |  |
| ≥3 | 2,506 | 2,305 (56.1) | 201 (70.8) |  | 1,416 | 1,368 (46.5) | 48 (59.4) |  | 1,090 | 937 (70.0) | 153 (74.6) |  |
| **Outcomes** |  |  |  |  |  |  |  |  |  |  |  |  |
| ICU admission | 991 | 931 (20.0) | 60 (21.6) | 0.66 | 646 | 627 (19.2) | 19 (17.7) | 0.75 | 345 | 304 (21.2) | 41 (22.9) | 0.72 |
| In-hospital death | 365 | 342 (8.4) | 23 (7.5) | 0.69 | 168 | 165 (5.7) | 3 (3.2) | 0.36 | 197 | 177 (12.3) | 20 (9.0) | 0.30 |
| Median length of stay in<br>days (IQR) |  | 4 (2–9) | 5 (3–8) | <.001 |  | 4 (2–7) | 4 (2–6) | <.001 |  | 6 (3–11) | 5 (3–9) | <.001 |

Abbreviations: BIPAP = bilevel positive airway pressure; CMO = comfort measures only; COVID-19 = coronavirus disease 2019; COVID-NET = COVID-19–Associated Hospitalization Surveillance Network; CPAP = continuous positive airway pressure; DNI = do not intubate; DNR = do not resuscitate; ICU = intensive care unit; IQR = interquartile range; LTCF = long-term care facility
<sup>a</sup>California, Connecticut, Colorado, Georgia, Maryland, Michigan, Minnesota, New Mexico, New York, Ohio, Oregon, Tennessee, and Utah
<sup>b</sup>Statistical significance for univariate analyses was determined as p<.10
<sup>c</sup>Race and Hispanic ethnicity: Data on race and ethnicity were categorized as follows: Hispanic or Latino, non-Hispanic White (White), non-Hispanic Black (Black), and Other/Unknown. If ethnicity was unknown (7% of cases), non-Hispanic ethnicity was assumed.
<sup>d</sup>Includes American Indian/Alaska Native (68, 1%), Asian/Pacific Islander (240, 7%), and multiple/unknown race (168, 3%). For adults aged <65 years, the breakdown was: American Indian/Alaska Native (50, 1%), Asian/Pacific Islander (164, 7%), and multiple/unknown race (117, 4%). For those aged ≥65 years, American Indian/Alaska Native (18, 1%), Asian/Pacific Islander (76, 7%), and multiple/unknown race (51, 2%).
<sup>e</sup>Long-term care facility (LTCF) residence was defined as residence in rehabilitation facilities, assisted living/residential care, group homes, nursing homes, skilled nursing facilities, long-term care facilities, long-term acute care hospitals, residential care facilities or other long-term care facilities.
<sup>f</sup>Obesity is defined as calculated body mass index (BMI) ≥30 kg/m<sup>2</sup>, and if BMI is missing, by International Classification of Diseases discharge diagnosis codes.
<sup>g</sup>Cardiovascular disease excludes hypertension.
<sup>h</sup>Overall condition categories as defined in Supplementary Table 1.

On multivariable analysis, after adjusting for, age, sex, race and Hispanic ethnicity, LTCF status, and underlying medical conditions, and excluding those whose reason for hospitalization was likely not primarily COVID-19-related, fully vaccinated patients aged ≥65 years, were more likely to be immunosuppressed (aRR: 3.2 (95% CI 2.0-5.1) and to be obese (aRR: 1.6 (95% CI 1.0-2.5) than unvaccinated patients **(Figure 1B)**. Fully vaccinated patients aged <65 years were more likely to have underlying neurologic and gastrointestinal/liver diseases (aRRs: 1.6 (95% CI 1.0-2.6) and 2.3 (95% CI 1.0-5.4), respectively).

**Figure 1.**
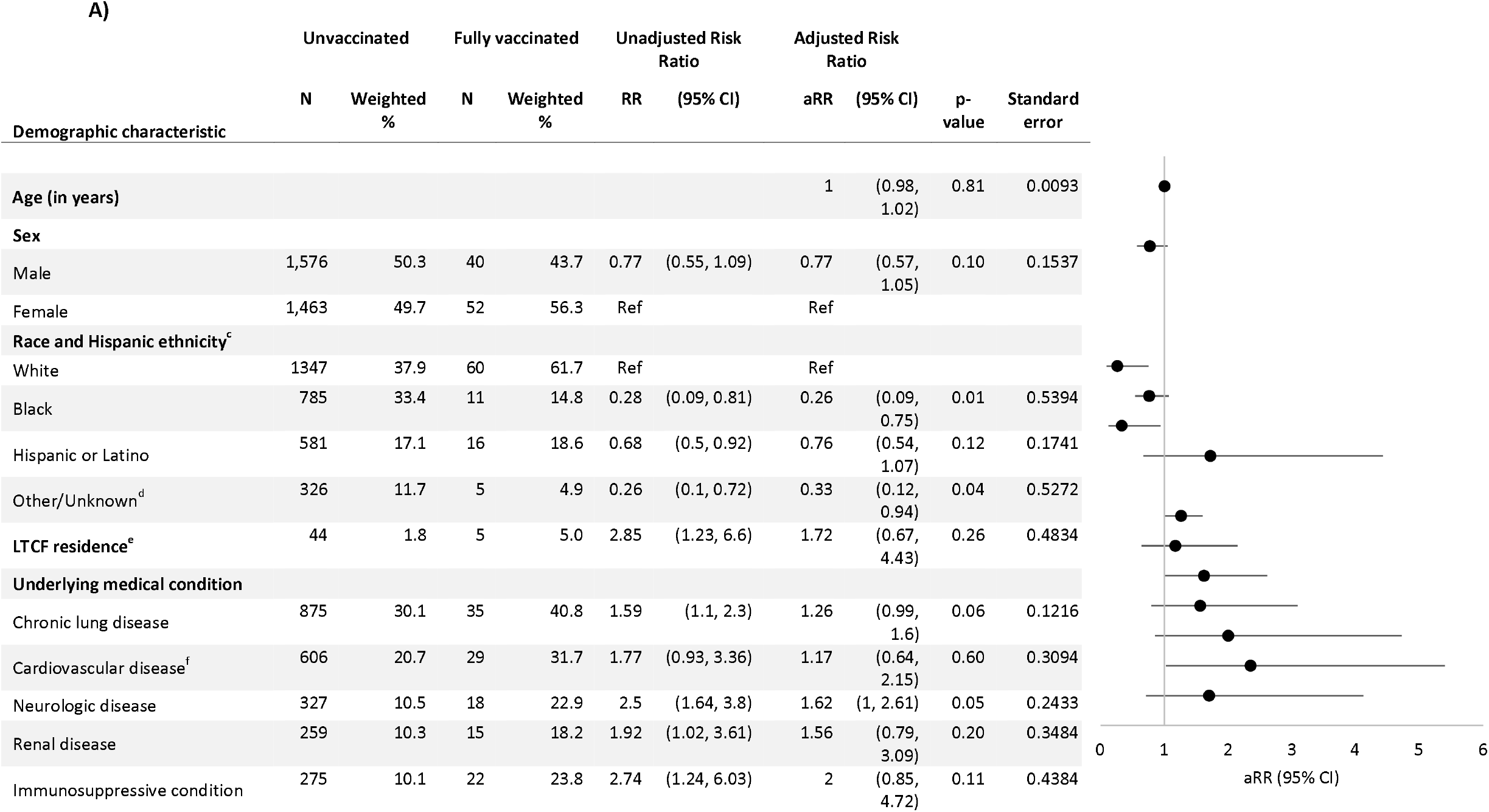
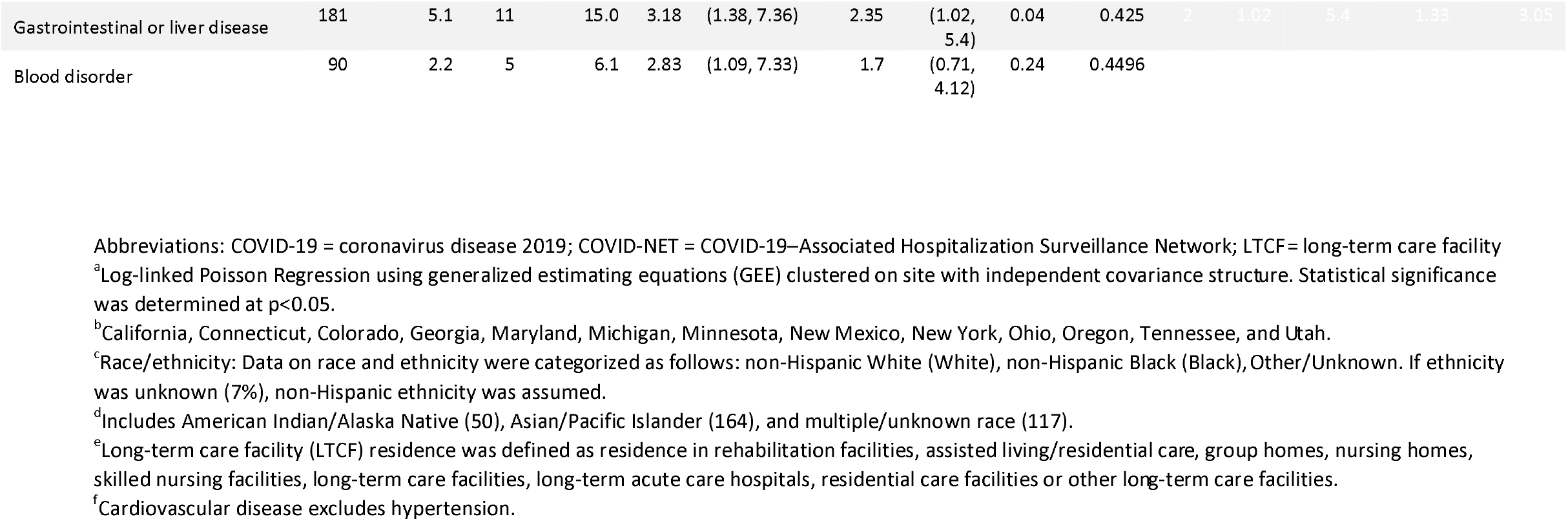
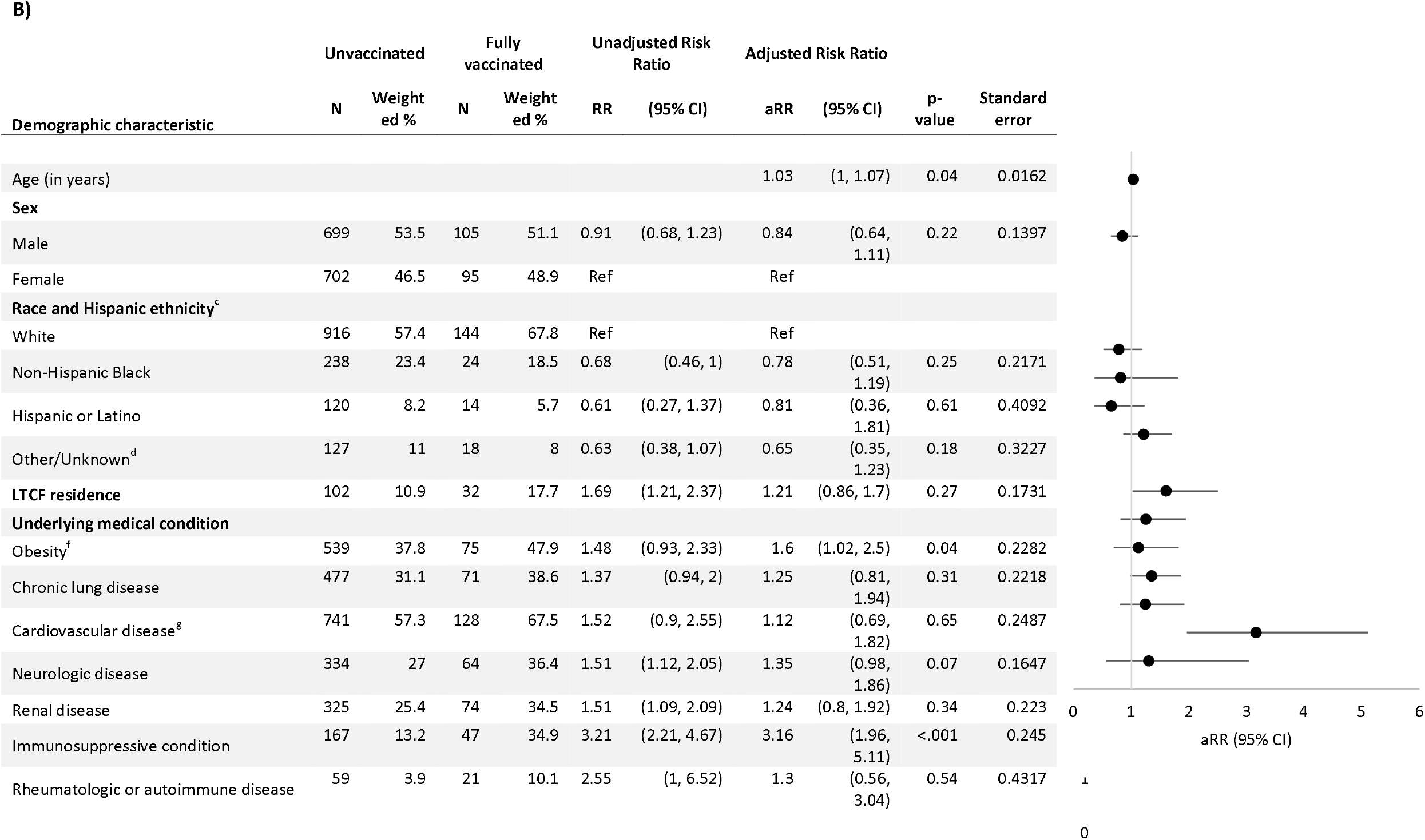
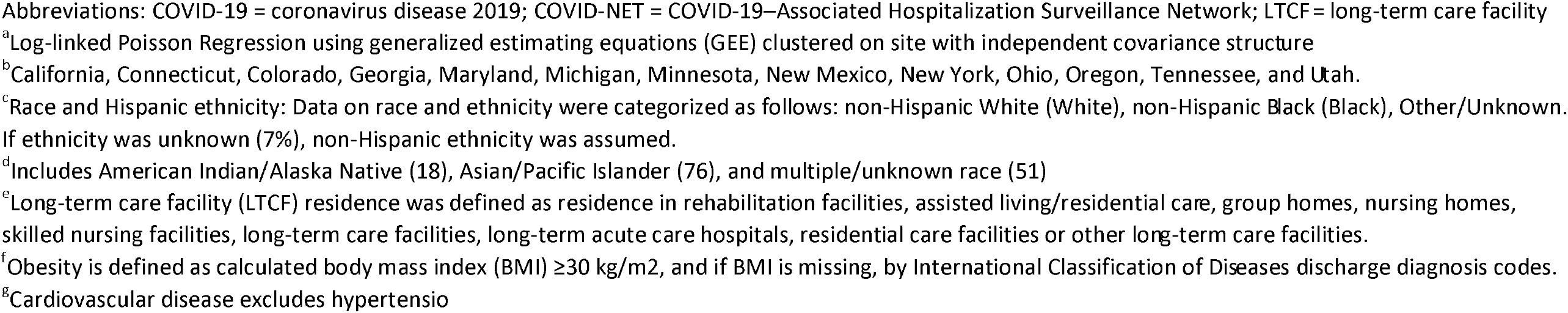
Multivariable model^a^ assessing factors associated with full vaccination status in hospitalized adults with laboratory-confirmed COVID-19-associated hospitalization admitted January 1 – June 30, 2021, restricting to those with COVID-19-associated hospitalization as the likely primary reason for admission, among those aged A) <65 years and B) aged ≥65 years — COVID-NET, 13 States^b^.

The number and proportion of fully vaccinated persons admitted to the ICU was similar to unvaccinated persons (60 (20.6%) v. 931 (20.0%), respectively; p-value=0.66), as were results for in-hospital death (23 (7.5%) v. 342 (8.4%), respectively; p-value=0.69). Median length of stay was significantly longer in fully vaccinated persons (median 5 days (IQR 3–8) v. 4 days (IQR 2–9), respectively; p<0.001) (**Table 2**). On multivariable analysis adjusting for age, sex, race, and underlying medical conditions, being fully vaccinated was significantly associated with reduced risk of severe disease (i.e. ICU admission and/or death) in those aged <65 years (aRR 0.8 (95% CI 0.6–1.0), but this association was not significant in those aged ≥65 years (aRR 1.1 (95% CI 0.7–1.6) (**Supplementary Tables 3A-B)**.

### Population-based rates of COVID-19-associated hospitalization by vaccination status

During the study period, the percentage of persons aged ≥18 years in the COVID-NET catchment area who were fully vaccinated increased from 0.9% in January to 57% in June 2021. The COVID-NET catchment population that was fully vaccinated varied by week and age group **(Supplementary Figure 3A–D)**. From January 24–July 24, 2021, the age-adjusted cumulative hospitalization rate among persons ≥18 years of age was 432 hospitalized cases per 100,000 unvaccinated persons compared with 26 per 100,000 fully vaccinated persons, a 17-fold higher cumulative rate **(Figure 2A)**; cumulative rate ratios were 26, 22 and 13 for those aged 18–49, 50–64, and ≥65 years respectively (**Supplementary Table 4)**. The weekly overall COVID-NET hospitalization rates tracked closely with the hospitalization rates of the unvaccinated population earlier in the study period, but as vaccination coverage increased, the overall rate started to converge with the rate in the fully vaccinated population (**Figure 2B, Figure 3A-C)**. The weekly rate ratio varied each week with no clear pattern (range in those aged ≥18 years: 6–31). For June 27–July 24, hospitalization rates were ≥10 times higher in unvaccinated persons compared with vaccinated persons for all age groups across all weeks (rate ratio range in those aged ≥18 years: 12-19) (**Supplementary Table 4)**.

**Figure 2.**
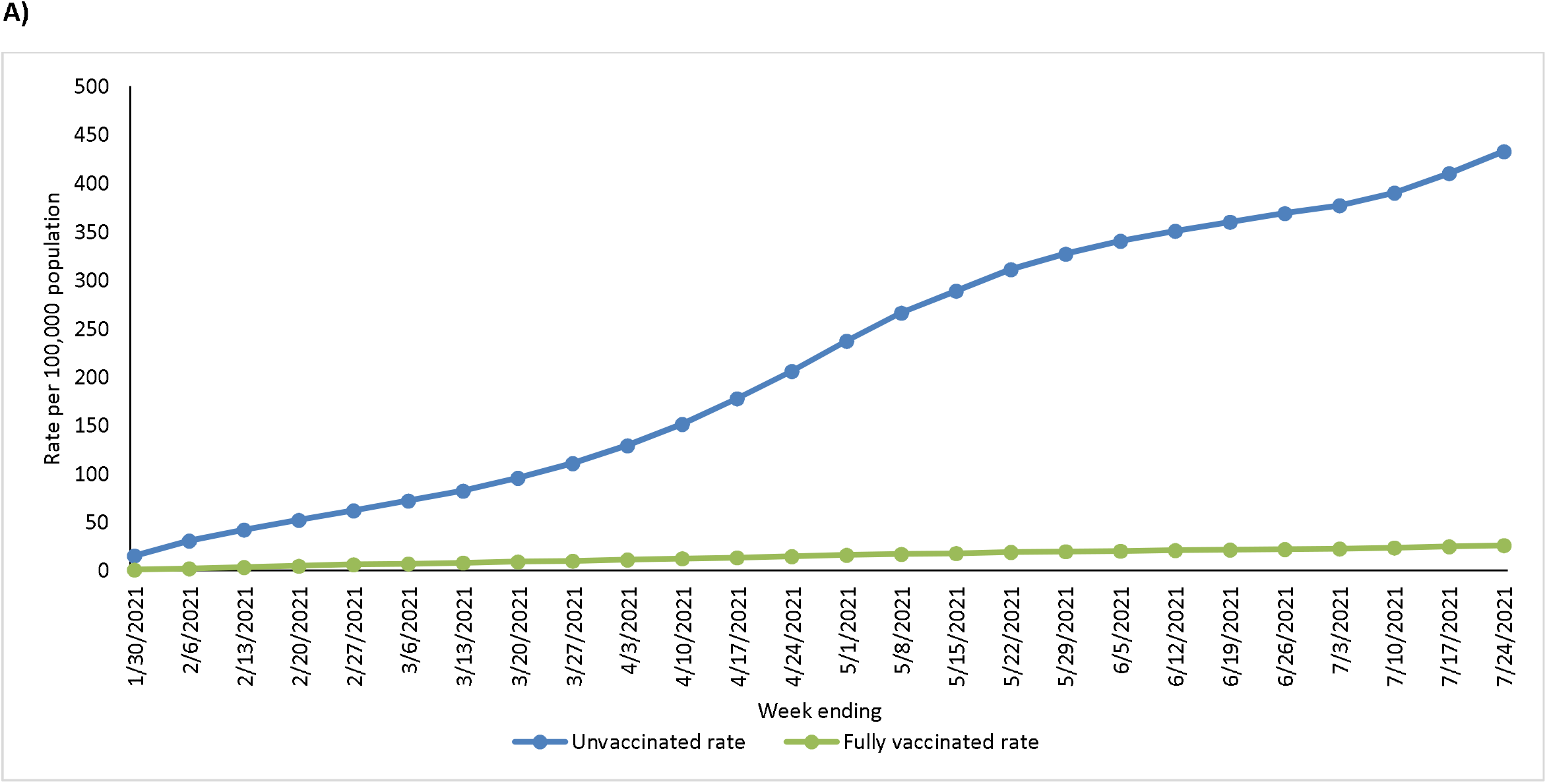
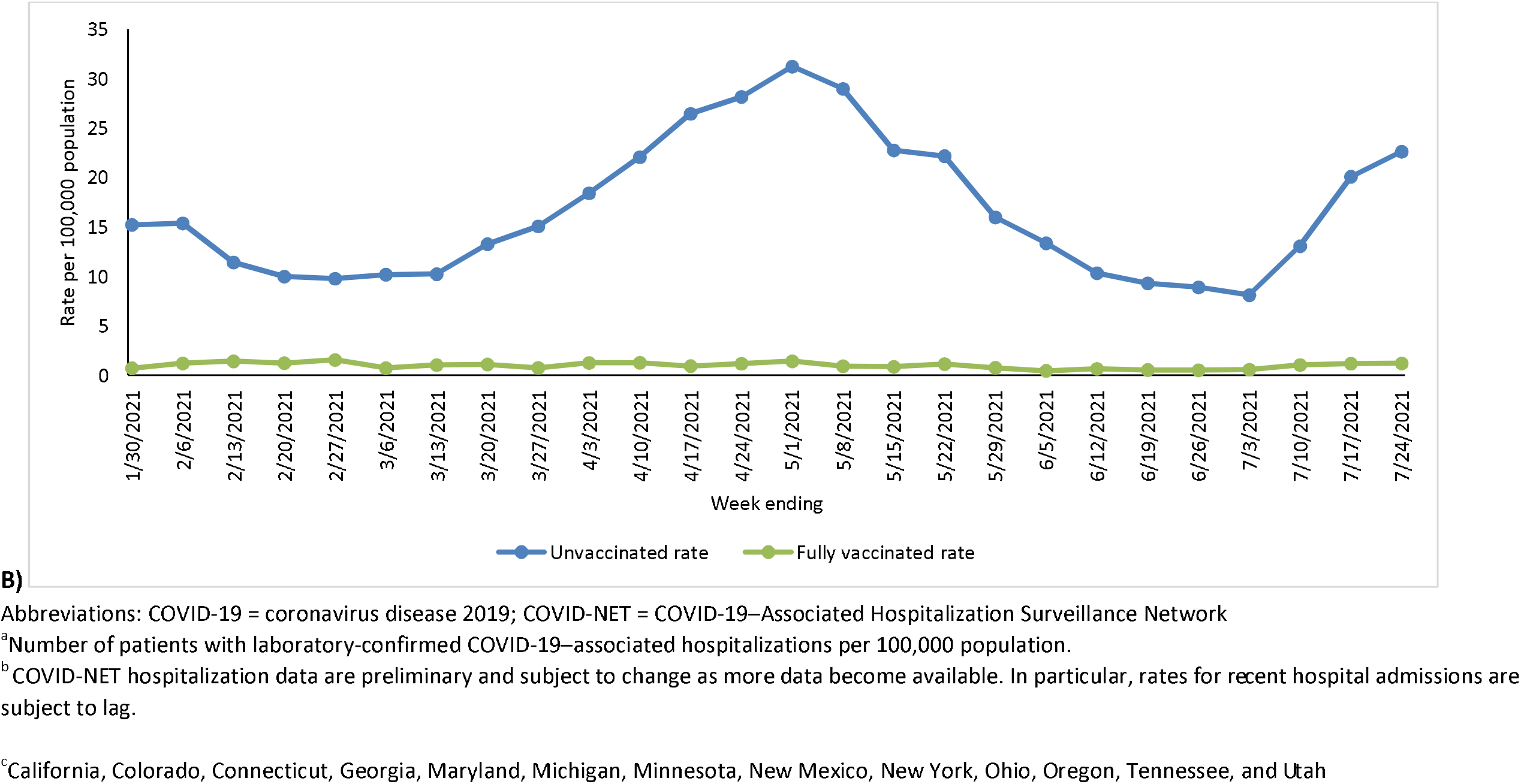
Age-adjusted population-based rates^a^ of COVID-19-associated hospitalizations among unvaccinated and fully vaccinated adults aged ≥18 years admitted January 24–July 24, 2021, ^b^ A) cumulative and B) by week of admission — COVID-NET, 13 States^c^.

**Figure 3.**
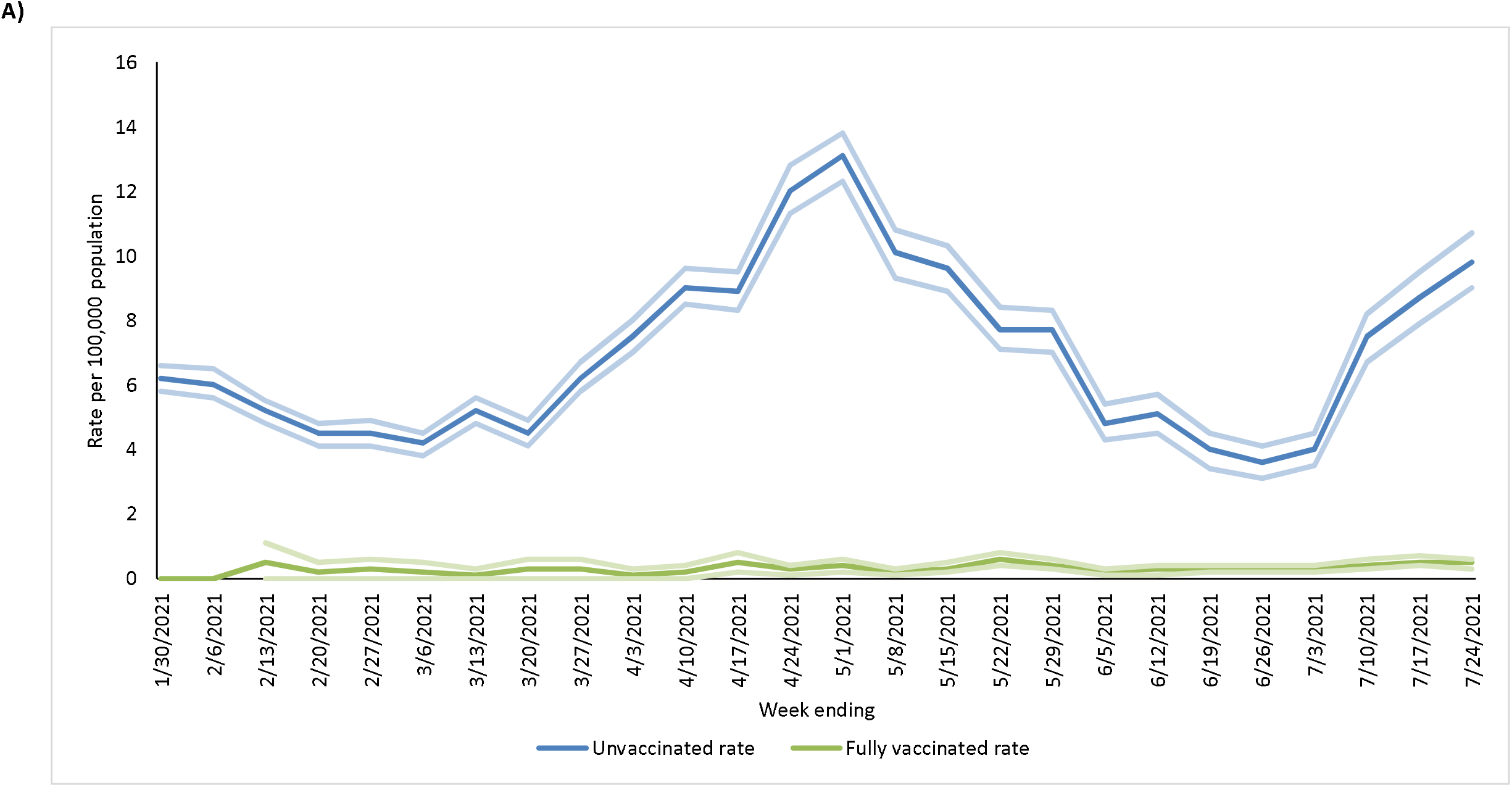
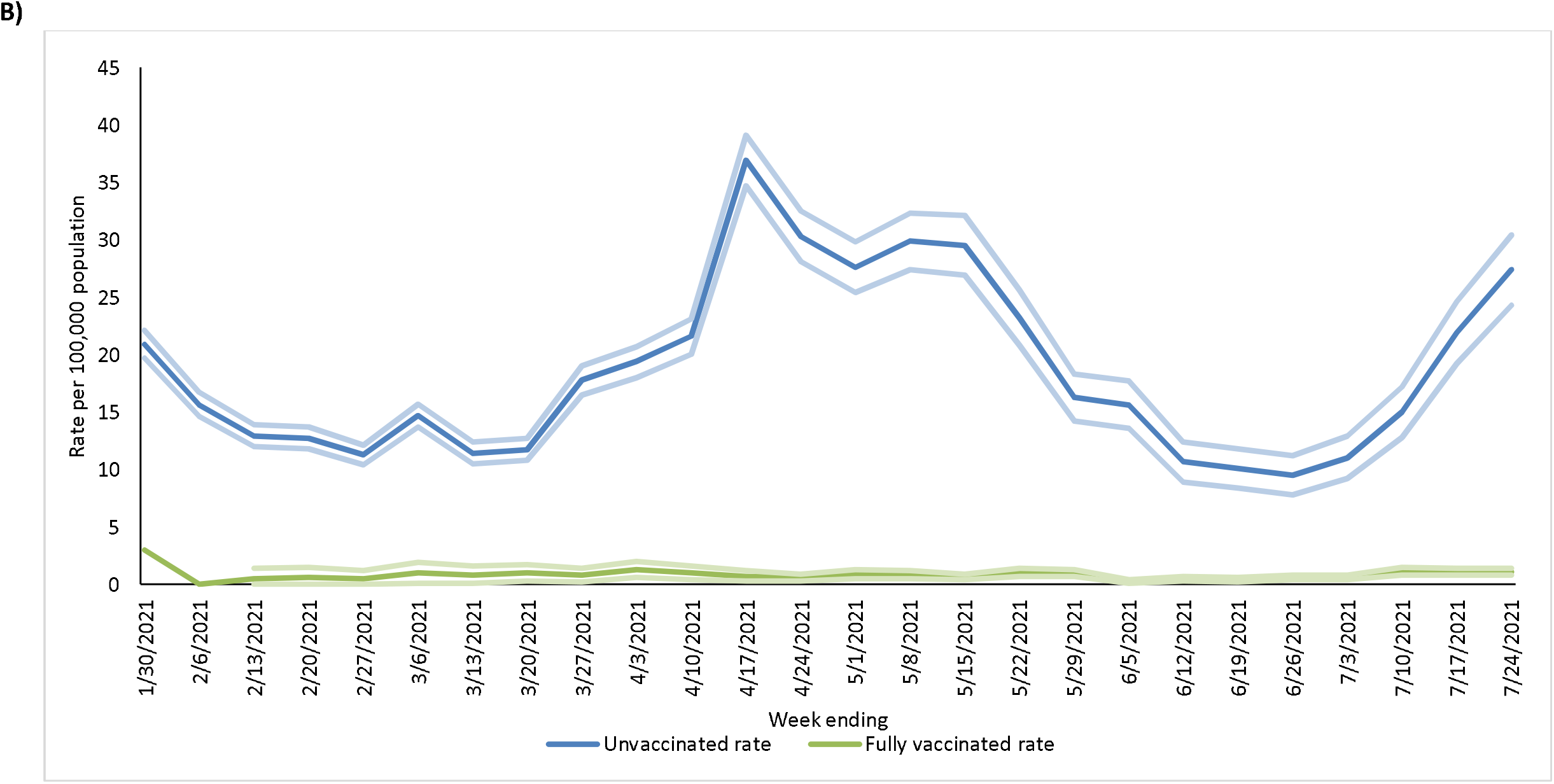
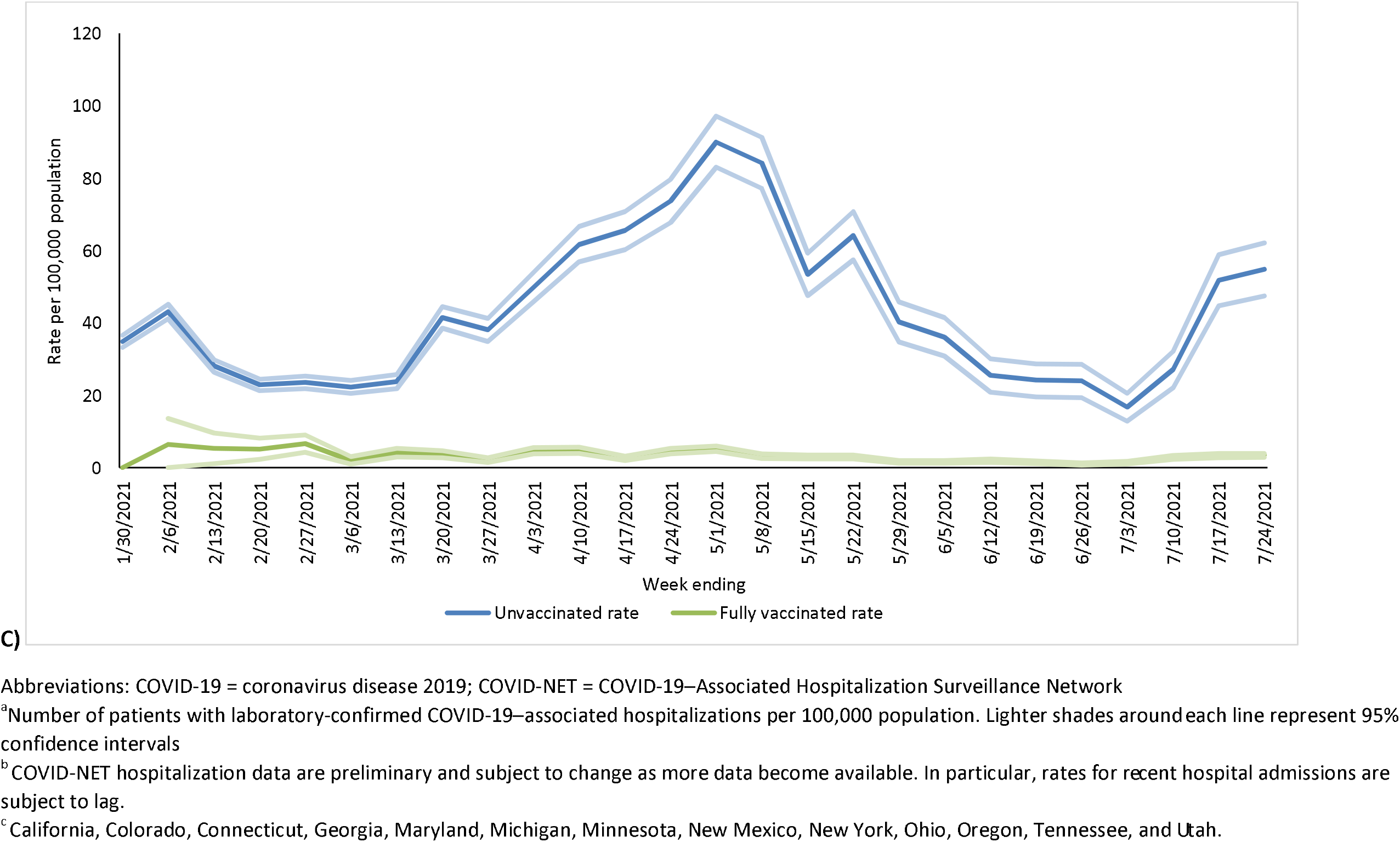
Age-specific population-based rates^a^ of COVID-19-associated hospitalizations among unvaccinated and fully vaccinated adults aged ≥18 years admitted January 24–July 24, 2021, ^b^ by week of admission and aged A) 18–49 years B) 50–64 years and C) ≥65 years — COVID-NET, 13 States^c^. Rates are shown with 95% confidence intervals.

## Discussion

In this analysis using data from a representative sample of over 67,000 laboratory-confirmed COVID-19-associated hospitalizations in persons aged ≥18 years, we demonstrated that cumulative population-based rates of COVID-19-associated hospitalization were approximately 17 times higher in unvaccinated persons compared with vaccinated persons, and more than 13 times higher in adults aged ≥65 years. Rates remained ≥10 times higher in unvaccinated persons compared with vaccinated persons in all adult age groups across all weeks in late June and July 2021, a period in which the Delta variant has predominated, suggesting that vaccines remain highly effective in preventing hospitalizations in all adult age groups. Those persons who were hospitalized despite being fully vaccinated were more medically fragile at baseline than those who were unvaccinated. Fully vaccinated cases were older, more likely to reside in LCTFs, and had more underlying medical conditions; nearly a third (32%) had immunosuppressive conditions, compared with 11% of those who were unvaccinated.

Our finding that a substantial and growing proportion of hospitalized persons were fully vaccinated is not surprising; the proportion of hospitalized cases who are fully vaccinated is expected to increase as population vaccination coverage increases. Given the high vaccination coverage, particularly in older age groups, which was more than 80% in July for those ≥65 years,^2^ the finding that 16% of hospitalized cases ≥18 years during that month were fully vaccinated is consistent with what we would expect from highly effective vaccines.

This analysis demonstrated that population-based rates of hospitalization were consistently far higher in unvaccinated persons compared with fully vaccinated persons in all age groups and over time. Using detailed IIS data from of a defined population, a 17-times greater cumulative hospitalization rate in unvaccinated persons is consistent with ongoing high levels of vaccine effectiveness in preventing hospitalizations and suggests that increasing vaccination coverage can prevent serious illness and death. Some of the differences in hospitalization rates between the unvaccinated and vaccinated populations may be attributable in part to differences in mask-wearing and physical distancing between the two groups. Cumulative rate ratios were lower in older adults than younger groups, but vaccinated adults aged ≥65 years had hospitalization rates that were more than 90% lower than unvaccinated persons in the same age group; recent increases in hospitalization rates appear to be driven disproportionately by unvaccinated persons. Far higher rates of hospitalization among unvaccinated persons compared to vaccinated persons of all ages persisted in July 2021, a time when the highly-transmissible Delta variant was the predominant variant circulating^6^ and hospitalization rates among vaccinated individuals remained comparatively low and stable. Although studies have suggested that current vaccines may be less effective in preventing SARS-CoV-2 infections due to the Delta variant compared than with previously circulating varians,^12-14^ this analysis suggests that vaccination remains highly effective against the Delta variant in preventing severe disease resulting in hospitalization.^6^

We demonstrated that, consistent with other studies, among hospitalized cases, vaccinated persons reflected older and more medically fragile populations that were prioritized to receive the vaccine early. Those aged ≥75 years, LTCF residents, those who are immunocompromised or have multiple underlying conditions may be among the most vulnerable to severe infection and the least likely to mount an adequate immune response to immunization; given early receipt of the vaccine, these populations may also be vulnerable to any potential waning of effectiveness over time.^4,15,16^ Increased vulnerability to severe disease resulting in hospitalization, even if fully vaccinated, supports the importance of vaccination among LCTF staff and visitors in protecting persons vulnerable to severe COVID-19. The high proportion of fully vaccinated patients who were immunocompromised also supports the recent recommendation for a third vaccine dose for immunocompromised adults who had previously received mRNA vaccines, although more research is needed to understand the extent to which an additional dose of vaccine results in increased protection in immunocompromised individuals.^17^

In the outpatient setting, vaccination likely attenuates disease severity if a breakthrough infection occurs,^5^ but our analysis showed that conditional on being hospitalized, vaccinated persons were still at a high risk of severe outcomes, with more than 20% admitted to the ICU. We did not find any clear difference in the risk for ICU admission or in-hospital death between vaccinated and unvaccinated persons. This may reflect the fact that those who were hospitalized despite vaccination may be more vulnerable to severe infection at baseline than those who are not vaccinated. In addition, unidentified confounders that are not well-accounted for may affect these results; further detailed analyses examining clinical presentation and outcomes are planned.

This analysis has several limitations. Although COVID-NET covers approximately 10% of the U.S. population, our findings may not be generalizable to the entire country. Since SARS-CoV-2 testing was conducted at the discretion of healthcare professionals, COVID-NET may not have captured all COVID-19-associated hospitalizations. The primary reason for hospital admission was not always clear, and some patients who met the COVID-NET case definition may have been hospitalized for reasons that were not primarily related to COVID-19. The inclusion of these hospitalizations in rate calculations could have resulted in an overestimation of hospitalization rates for COVID-19 for both unvaccinated and vaccinated persons.

Using population-based hospitalization rates, our study found unvaccinated adults are hospitalized at rates that are 17 times higher than for vaccinated adults in the same catchment areas. Rates of COVID-19-associated hospitalizations are far higher in unvaccinated persons in all adult age groups, even during a period when the highly-transmissible Delta variant was the predominant circulating variant.^12^ Population-based hospitalization rates will be a critical metric in the continued assessment of the effectiveness of vaccines in the population, and complementary to vaccine effectiveness studies using other methods.^4,18^ The much higher rates observed in unvaccinated persons compared to vaccinated adults suggest that vaccines remain highly effective in preventing hospitalization for COVID-19 illness. Continuing to increase vaccination coverage in all eligible groups is crucial to preventing severe COVID-19.

## Data Availability

Publicly available data referred to in this analysis can be found at: https://gis.cdc.gov/grasp/covidnet/covid19_3.html
https://gis.cdc.gov/grasp/COVIDNet/COVID19_5.html

https://gis.cdc.gov/grasp/COVIDNet/COVID19_5.html

## Disclaimer

The findings and conclusions in this report are those of the authors and do not necessarily represent the views of the U.S. Centers for Disease Control and Prevention.

## Acknowledgments

Roxanne Archer, Susan Brooks, Monica Napoles, Jeremy Roland, Tiffany Tsukuda, CA Emerging Infections Program; Arthur Reingold, University of California, Berkeley; Rachel Herlihy, Breanna Kawasaki, Colorado Department of Public Health and Environment; Ann Basting, Tessa Carter, Maria Correa, Daewi Kim, Carol Lyons, Amber Maslar, Julie Plano, Connecticut Emerging Infections Program, Yale School of Public Health; Katelyn Ward, Georgia Emerging Infections Program, Georgia Department of Health, Division of Infectious Diseases, School of Medicine, Emory University; Marina Bruck, Rayna Ceasar, Gracie Chambers, Taylor Eisenstein, Emily Fawcett, Asmith Joseph, Grayson Kallas, Stephanie Lehman, Jana Manning, Annabel Patterson, Allison Roebling, Chandler Surell, Georgia Emerging Infections Program, Georgia Department of Health, Veterans Affairs Medical Center, Foundation for Atlanta Veterans Education and Research; David Blythe, Alicia Brooks, Elisabeth Vaeth, Cindy Zerrlaut, Maryland Department of Health; Rachel Park, Michelle Wilson, Maryland Emerging Infections Program - The Johns Hopkins Bloomberg School of Public Health; Chloe Brown, Jim Collins, Justin Henderson, Shannon Johnson, Sue Kim, Lauren Leegwater, Sierra Peguies-Khan, Val Tellez Nunez, Michigan Department of Health and Human Services; Aaron Bieringer, Erica Bye, Richard Danila, Kristen Ehresmann, Corinne Holtzman, Sydney Kuramoto, Stephanie Meyer, Miriam Muscoplat, Rachel Ostadkar, Minnesota Department of Health; Marianne Murphy, Murtada Khalifa, Yassir Talha, CDC Foundation/New Mexico Department of Health; Melissa Judson, Sunshine Martinez, Jasmyn Sanchez, Chad Smelser, Daniel Sosin, New Mexico Department of Health; Kathy M. Angeles, Melissa Christian, Nancy Eisenberg, Emily B. Hancock, Sarah A. Khanlian, Sarah Lathrop, Wickliffe Omondi, Mayvilynne Poblete, Dominic Rudin, Yadira Salazar-Sanchez, Sarah Shrum Davis, Chelsea McMullen, Susan Ropp, New Mexico Emerging Infections Program; Kerianne Engesser, Suzanne McGuire, Adam Rowe, Nancy Spina, New York State Department of Health; Nancy Bennett, Virginia Cafferky, Maria Gaitan, Christine Long, Thomas Peer, Kevin Popham, University of Rochester School of Medicine and Dentistry; Kylie Seeley, Oregon Health & Science University School of Medicine; Ama Owusu-Dommey, Emily Youngers, Breanna McArdle, Public Health Division; Oregon Health Authority; Kathy Billings, Katie Dyer, Anise Elie, Karen Leib, Tiffanie Markus, Terri McMinn, Danielle Ndi, Manideepthi Pemmaraju, John Ujwok, Vanderbilt University Medical Center; Ian Buchta, Amanda Carter, Ryan Chatelain, Andrew Haraghey, Laine McCullough, Jacob Ortega, Andrea Price, Tyler Riedesel, Ilene Risk, Caitlin Shaw, Melanie Spencer, Ashley Swain, Salt Lake County Health Department; Keegan McCaffrey, Utah Department of Health; Kelly Oakeson, Alessandro Rossi, Utah Public Health Laboratory

## Figures

**Figure 1. Multivariable model^a^ assessing factors associated with full vaccination status in hospitalized adults with laboratory-confirmed COVID-19-associated hospitalization admitted January 1–June 30, 2021, restricting to those with COVID-19-associated hospitalization as the likely primary reason for admission, among those aged A) <65 years and B) ≥65 years — COVID-NET, 13 States^b^**

**Figure 2. Age-adjusted population-based rate^a^ of COVID-19-associated hospitalizations among unvaccinated and fully vaccinated adults ≥18 years of age admitted January 24–June 26, 2021, A) cumulative and B) by week of admission — COVID-NET, 13 States^b^**

**Figure 3. Age-specific population-based rates^a^ of COVID-19-associated hospitalizations among unvaccinated and fully vaccinated adults ≥18 years of age admitted January 24–July 24, 2021, by week of admission and age A) 18–49 years B) 50–64 years and C) ≥65 years — COVID-NET, 13 States^b^**

## Supplementary material

**Supplementary Table 1. Categorization of Underlying Medical Conditions ^a^**

**Supplementary Table 2. Characteristics of fully vaccinated cases who had a prior positive SARS-CoV-2 test compared with fully vaccinated cases with no evidence of prior infection, among adults ≥ 18 years admitted January 1–June 30, 2021 — COVID-NET, 13 States^a^**

**Supplementary Table 3A. Multivariable model^a^ assessing factors associated with severe COVID-19 diseases (ICU, in-hospital death) in hospitalized adults with laboratory-confirmed COVID-19-associated hospitalization admitted January 1 – June 30, 2021, restricting to those with COVID-19-associated hospitalization as the likely primary reason for admission, among those aged <65 years — COVID-NET, 13 States^b^**

**Supplementary Table 3B. Multivariable model^a^ assessing factors associated with severe COVID-19 diseases (ICU, in-hospital death) in hospitalized adults with laboratory-confirmed COVID-19-associated hospitalization admitted January 1–June 30, 2021, restricting to those with COVID-19-associated hospitalization as the likely primary reason for admission, among those aged ≥65 years — COVID-NET, 13 States^b^**

**Supplementary Table 4. Rate ratios of COVID-19-associated hospitalizations in unvaccinated vs vaccinated patients among all adults aged ≥18 years and by age group^a^, January 24–June 26, 2021^b^ — COVID-NET, 13 States^c^**

## Supplementary Figures

**Supplementary Figure 1. Selection of cases for analysis of adults ≥18 years with laboratory-confirmed COVID-19-associated hospitalization admitted January 1–June 30, 2021, by vaccination status — COVID-NET, 13 States^a^**

**Supplementary Figure 2. Proportion of adults ≥18 years with COVID-19-associated hospitalizations admitted January 1 – June 30, 2021 who are fully vaccinated, by age group and month of admission — COVID-NET, 13 States**

**Supplementary Figure 3. COVID-NET catchment population by vaccination status and week, used as denominator in population-based-rate calculations^a^ among A) all adults aged ≥18 years, B) 18–49 years, C) 50–64 years, and D) ≥65 years — COVID-NET^c^, January 24–July 24, 2021— COVID-NET, 13 States**

## Supplementary Content

### Supplementary Tables

**Supplement Table 1.** Categorization of Underlying Medical Conditions^a^.

|  |  |
| --- | --- |
| Chronic Lung Disease | • Active tuberculosis<br>• Asbestosis<br>• Asthma/Reactive airway disease<br>• Bronchiectasis<br>• Bronchiolitis obliterans<br>• Chronic bronchitis<br>• Chronic respiratory failure<br>• Cystic fibrosis<br>• Emphysema/Chronic Obstructive Pulmonary Disease<br>• Interstitial lung disease<br>• Obstructive sleep apnea<br>• Oxygen dependent<br>• Pulmonary fibrosis<br>• Restrictive lung disease<br>• Sarcoidosis |
| Diabetes Mellitus |  |
| Other Chronic Metabolic Disease (except Diabetes Mellitus) | • Adrenal Disorders (Addison's, Adrenal Insufficiency, Cushing Syndrome, Congenital Adrenal Hyperplasia)<br>• Glycogen or other storage diseases<br>• Hyper/Hypo function of pituitary gland<br>• Inborn errors of metabolism<br>• Metabolic Syndrome<br>• Parathyroid Dysfunction (Hyperparathyroidism, Hypoparathyroidism)<br>• Thyroid dysfunction (Grave's disease, Hashimoto's Disease, Hyperthyroidism, Hypothyroidism) |
| Blood Disorders/Hematologic Conditions | • Alpha thalassemia<br>• Aplastic anemia<br>• Beta thalassemia<br>• Coagulopathy (Factor V Leiden, Von Willebrand Disease)<br>• Hemoglobin S-beta thalassemia<br>• Leukopenia<br>• Myelodysplastic syndrome<br>• Neutropenia<br>• Pancytopenia<br>• Polycythemia vera<br>• Sickle cell disease<br>• Splenectomy/Asplenia<br>• Thrombocytopenia |
| Cardiovascular Disease (except hypertension) | • Aortic aneurysm, history of<br>• Aortic/Mitral/Tricuspid/Pulmonic valve replacement, history of<br>• Aortic regurgitation<br>• Aortic stenosis<br>• Atherosclerotic cardiovascular disease<br>• Atrial fibrillation |
|  | • Atrioventricular blocks<br>• Automated implantable devices/Pacemaker<br>• Bundle branch block<br>• Cardiomyopathy<br>• Carotid stenosis<br>• Cerebral vascular accident/Incident/Stroke, history of<br>• Congenital heart disease<br>• Coronary artery bypass grafting, history of<br>• Coronary artery disease<br>• Deep vein thrombosis, history of<br>• Heart failure/congestive heart failure<br>• Myocardial infarction, history of<br>• Mitral regurgitation<br>• Mitral stenosis<br>• Peripheral artery disease<br>• Peripheral vascular disease<br>• Pulmonary embolism, history of<br>• Pulmonary hypertension<br>• Pulmonic regurgitation<br>• Pulmonic stenosis<br>• Transient ischemic attack, history of<br>• Tricuspid regurgitation<br>• Tricuspid stenosis<br>• Ventricular fibrillation, history of<br>• Ventricular tachycardia, history of |
| Hypertension |  |
| Neurologic Disorder | • Amyotrophic lateral sclerosis<br>• Cerebral palsy<br>• Cognitive dysfunction<br>• Dementia/Alzheimer's disease<br>• Developmental delay<br>• Down syndrome/Trisomy 21<br>• Edward's syndrome/Trisomy 18<br>• Epilepsy/seizure/seizure disorder<br>• Mitochondrial disorder<br>• Multiple sclerosis<br>• Muscular dystrophy<br>• Myasthenia gravis<br>• Neural tube defects/Spina bifida<br>• Neuropathy<br>• Parkinson's disease<br>• Plegias/Paralysis/Quadriplegia<br>• Scoliosis/Kyphoscoliosis<br>• Traumatic brain injury, history of |
| History of Guillain-Barre Syndrome |  |
| Immunocompromised Conditions | • AIDS or CD4 count <200<br>• Complement deficiency<br>• Graft vs. host disease<br>• HIV Infection<br>• Immunoglobulin deficiency/Immunodeficiency |
|  | • Immunosuppressive therapy (within the last 12 months prior to admission)<br>• Leukemia<sup>b</sup><br>• Lymphoma/Hodgkins/Non-Hodgkins<sup>b</sup><br>• Metastatic cancer<sup>b</sup><br>• Multiple myeloma<sup>b</sup><br>• Solid organ malignancy<sup>b</sup><br>• Steroid therapy (within 2 weeks of admission)<br>• Transplant, hematopoietic stem cell (Bone marrow transplant, peripheral stem cell transplant), history of<br>• Transplant, solid organ, history of |
| Renal Disease | • Chronic kidney disease/chronic renal insufficiency<br>• Dialysis<br>• End stage renal disease<br>• Glomerulonephritis<br>• Nephrotic syndrome<br>• Polycystic kidney disease |
| Gastrointestinal/Liver Disease | • Alcoholic hepatitis<br>• Autoimmune hepatitis<br>• Barrett's esophagitis<br>• Chronic liver disease<br>• Chronic pancreatitis<br>• Cirrhosis/End stage liver disease<br>• Crohn's disease<br>• Esophageal varices<br>• Esophageal strictures<br>• Hepatitis B, chronic<br>• Hepatitis C, chronic<br>• Non-alcoholic fatty liver disease/Non-alcoholic steatohepatitis<br>• Ulcerative colitis |
| Rheumatologic/Autoimmune Conditions | • Ankylosing spondylitis<br>• Dermatomyositis<br>• Juvenile idiopathic arthritis<br>• Kawasaki disease<br>• Microscopic polyangiitis<br>• Polyarteritis nodosum<br>• Polymyalgia rheumatica<br>• Polymyositis<br>• Psoriatic arthritis<br>• Rheumatoid arthritis<br>• Systemic Lupus Erythematosus/Lupus<br>• Systemic sclerosis<br>• Takayasu arteritis<br>• Temporal/Giant Cell arteritis<br>• Vasculitis, other |
| Other, specify | • Post-partum (2 weeks or less)<br>• Feeding tube dependent<br>• Tracheostomy dependent/ventilator dependent<br>• Wheelchair dependent<br>• Free text field for other underlying medical conditions that |
|  | were not specified above |

<sup>a</sup>Underlying medical conditions were selected before hospitalization surveillance began.
<sup>b</sup>Current/in treatment or diagnosed in previous 12 months

**Supplementary Table 2.** Characteristics of fully vaccinated cases who had a prior positive SARS-CoV-2 test compared with fully vaccinated cases with no evidence of prior infection, among adults ≥ 18 years admitted January 1–June 30, 2021 — COVID-NET, 13 States^a^.

| Characteristics | No evidence of prior infection<br>n=292 | Prior positive test<br>n=47<sup>b</sup> | p-value<sup>c</sup> |
| --- | --- | --- | --- |
| **Age group** |  |  | 0.82 |
| 18-49 years | 29 (8.7) | 4 (5.9) |  |
| 50-64 years | 63 (16.3) | 11 (18.6) |  |
| ≥65 years | 200 (75.0) | 32 (75.5) |  |
| **Sex** |  |  | 0.77 |
| Male | 145 (49.3) | 26 (45.4) |  |
| Female | 147 (50.7) | 21 (54.6) |  |
| **Race and Hispanic ethnicity<sup>d</sup>** |  |  |  |
| White | 204 (66.3) | 33 (69.9) | 0.10 |
| Black | 35 (17.5) | 7 (15.5) |  |
| Hispanic or Latino | 30 (9.0) | 4 (9.4) |  |
| Other/Unknown | 23 (7.2) | 3 (5.2) |  |
| **Month of admission** |  |  | **0.08** |
| January | 1 (0.6) | 0 (0.0) |  |
| February | 2 (1.6) | 0 (0.0) |  |
| March | 18 (13.0) | 5 (37.1) |  |
| April | 39 (34.2) | 7 (21.8) |  |
| May | 99 (31.2) | 13 (22.7) |  |
| June | 133 (19.3) | 22 (18.4) |  |
| **Code status on admission** |  |  | 0.62 |
| Full code | 222 (74.5) | 36 (82.5) |  |
| DNR/DNI/CMO | 47 (17.3) | 9 (10.9) |  |
| Unknown | 23 (8.2) | 2 (6.6) |  |
| **LTCH residence<sup>e</sup>** | 37 (14.5) | 16 (31.6) | **0.07** |
| **Any symptom** | 250 (88.2) | 35 (65.0) | **0.02** |
| **Hypertension** | 202 (70.5) | 33 (75.6) | 0.61 |
| **Obesity<sup>f</sup>** | 121 (48.4) | 15 (46.5) | 0.90 |
| **Severe obesity<sup>g</sup>** | 34 (14.1) | 5 (8.9) | 0.39 |
| **Chronic metabolic disease** | 142 (48.5) | 23 (63.6) | 0.20 |
| Diabetes mellitus | 106 (38.1) | 18 (53.5) | 0.23 |
| **Chronic lung disease** | 106 (39.2) | 23 (43.7) | 0.72 |
| Asthma/Reactive airway disease | 46 (18.7) | 10 (18.8) | 0.99 |
| COPD | 43 (14.2) | 11 (21.7) | 0.36 |
| **Cardiovascular disease<sup>h</sup>** | 157 (58.6) | 30 (78.0) | 0.04 |
| Heart failure | 57 (24.6) | 17 (34.1) | 0.40 |
| Coronary artery disease | 58 (18.6) | 12 (19.1) | 0.94 |
| **Neurologic disease** | 82 (33.0) | 18 (31.9) | 0.92 |
| **Renal disease** | 89 (30.4) | 14 (21.2) | 0.32 |
| **Immunosuppressive condition** | 69 (32.1) | 6 (9.8) | **0.006** |
| **Gastrointestinal or liver disease** | 24 (7.3) | 6 (9.6) | 0.63 |
| **Blood disorder** | 19 (5.4) | 4 (2.8) | 0.30 |
| **Rheumatologic or autoimmune disease** | 24 (8.7) | 1 (3.0) | 0.28 |
| **Number of underlying conditions<sup>i</sup>** |  |  | **0.07** |
| 0 | 13 (3.6) | 0 (0.0) |  |
| 1 | 24 (6.3) | 9 (17.3) |  |
| 2 | 54 (19.3) | 5 (5.1) |  |
| ≥3 | 201 (70.8) | 33 (77.7) |  |
| **ICU admission** | 60 (21.6) | 7 (11.4) | 0.22 |
| **In-hospital death** | 23 (7.5) | 2 (1.2) | **0.006** |
| **ICU admission or in-hospital death** | 66 (23.3) | 8 (12.0) | 0.18 |
| **Median length of stay in days (IQR)** | 5 (3–8) | 5 (2–14) | **<.001** |
| **ICU admission, invasive mechanical ventilation, or in-hospital death** | 69 (23.9) | 10 (34.3) | 0.47 |
| **Invasive mechanical ventilation** | 25 (8.7) | 3 (23.0) | 0.23 |
| **Vasopressor use** | 29 (11.0) | 5 (4.9) | 0.19 |
| **BIPAP or CPAP use** | 44 (14.5) | 4 (6.7) | 0.31 |
| **Renal Replacement Therapy** | 14 (3.1) | 2 (1.2) | 0.23 |

Abbreviations: BIPAP = bilevel positive airway pressure; CMO = comfort measures only; COVID-NET = COVID-19–Associated Hospitalization Surveillance Network; CPAP = continuous positive airway pressure; DNI = do not intubate; DNR = do not resuscitate; ICU = intensive care unit; IQR = interquartile range; LTCF = long-term care facility; SARS-CoV-2 = severe acute respiratory syndrome coronavirus 2
<sup>a</sup>California, Connecticut, Colorado, Georgia, Maryland, Michigan, Minnesota, New Mexico, New York, Ohio, Oregon, Tennessee, and Utah
<sup>b</sup>Five cases with hospitalization not primarily associated with COVID-19 illness were excluded
<sup>c</sup>Statistical significance for univariate analyses was determined as $p < .10$
<sup>d</sup>Race and Hispanic ethnicity: Data on race and ethnicity were categorized as follows: Hispanic or Latino, non-Hispanic White (White), non-Hispanic Black (Black), and Other/Unknown. If ethnicity was unknown, non-Hispanic ethnicity was assumed.
<sup>e</sup>Long-term care facility (LTCF) residence was defined as residence in rehabilitation facilities, assisted living/residential care, group homes, nursing homes, skilled nursing facilities, long-term care facilities, long-term acute care hospitals, residential care facilities or other long-term care facilities.
<sup>f</sup>Obesity is defined as calculated body mass index (BMI) $\geq 30$ kg/m<sup>2</sup>, and if BMI is missing, by International Classification of Diseases discharge diagnosis codes.
<sup>g</sup>Severe obesity is defined as calculated body mass index (BMI) $\geq 40$ kg/m<sup>2</sup>, and if BMI is missing, by International Classification of Diseases discharge diagnosis codes.
<sup>h</sup>Cardiovascular disease excludes hypertension
<sup>i</sup>Overall condition categories as defined in Supplementary Table 1

**Supplementary Table 3A.**
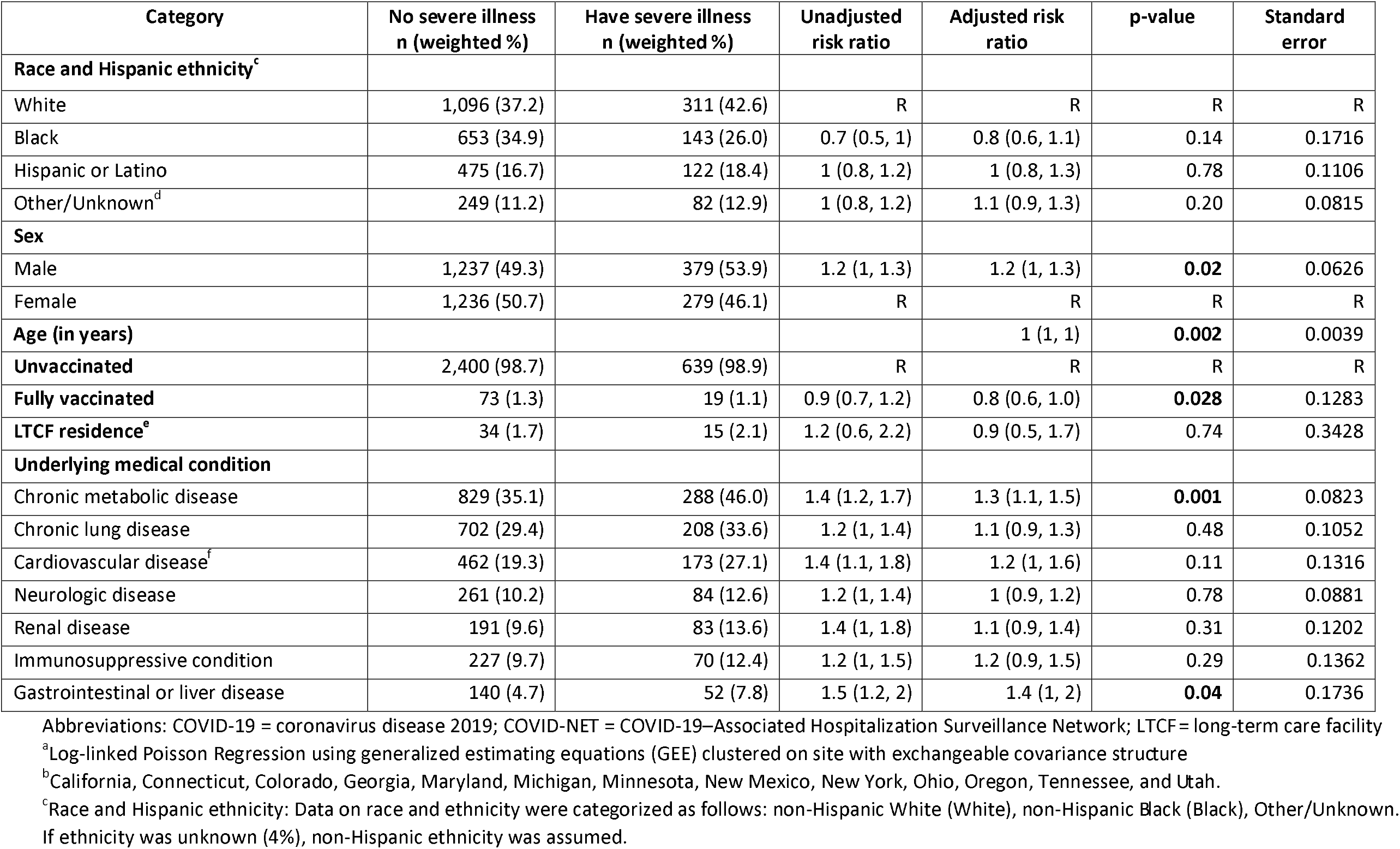
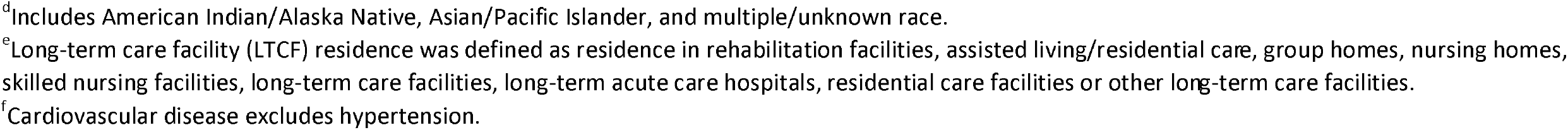
Multivariable model^a^ assessing factors associated with severe COVID-19 diseases (ICU, in-hospital death) in hospitalized adults with laboratory-confirmed COVID-19-associated hospitalization admitted January 1 – June 30, 2021, restricting to those with COVID-19-associated hospitalization as the likely primary reason for admission, among those aged <65 years — COVID-NET, 13 States^b^.

| Category | No severe illness<br>n (weighted %) | Have severe illness<br>n (weighted %) | Unadjusted<br>risk ratio | Adjusted risk<br>ratio | p-value | Standard<br>error |
| --- | --- | --- | --- | --- | --- | --- |
| **Race and Hispanic ethnicity<sup>c</sup>** |  |  |  |  |  |  |
| White | 1,096 (37.2) | 311 (42.6) | R | R | R | R |
| Black | 653 (34.9) | 143 (26.0) | 0.7 (0.5, 1) | 0.8 (0.6, 1.1) | 0.14 | 0.1716 |
| Hispanic or Latino | 475 (16.7) | 122 (18.4) | 1 (0.8, 1.2) | 1 (0.8, 1.3) | 0.78 | 0.1106 |
| Other/Unknown<sup>d</sup> | 249 (11.2) | 82 (12.9) | 1 (0.8, 1.2) | 1.1 (0.9, 1.3) | 0.20 | 0.0815 |
| **Sex** |  |  |  |  |  |  |
| Male | 1,237 (49.3) | 379 (53.9) | 1.2 (1, 1.3) | 1.2 (1, 1.3) | **0.02** | 0.0626 |
| Female | 1,236 (50.7) | 279 (46.1) | R | R | R | R |
| **Age (in years)** |  |  |  | 1 (1, 1) | **0.002** | 0.0039 |
| **Unvaccinated** | 2,400 (98.7) | 639 (98.9) | R | R | R | R |
| **Fully vaccinated** | 73 (1.3) | 19 (1.1) | 0.9 (0.7, 1.2) | 0.8 (0.6, 1.0) | **0.028** | 0.1283 |
| **LTCF residence<sup>e</sup>** | 34 (1.7) | 15 (2.1) | 1.2 (0.6, 2.2) | 0.9 (0.5, 1.7) | 0.74 | 0.3428 |
| **Underlying medical condition** |  |  |  |  |  |  |
| Chronic metabolic disease | 829 (35.1) | 288 (46.0) | 1.4 (1.2, 1.7) | 1.3 (1.1, 1.5) | **0.001** | 0.0823 |
| Chronic lung disease | 702 (29.4) | 208 (33.6) | 1.2 (1, 1.4) | 1.1 (0.9, 1.3) | 0.48 | 0.1052 |
| Cardiovascular disease<sup>f</sup> | 462 (19.3) | 173 (27.1) | 1.4 (1.1, 1.8) | 1.2 (1, 1.6) | 0.11 | 0.1316 |
| Neurologic disease | 261 (10.2) | 84 (12.6) | 1.2 (1, 1.4) | 1 (0.9, 1.2) | 0.78 | 0.0881 |
| Renal disease | 191 (9.6) | 83 (13.6) | 1.4 (1, 1.8) | 1.1 (0.9, 1.4) | 0.31 | 0.1202 |
| Immunosuppressive condition | 227 (9.7) | 70 (12.4) | 1.2 (1, 1.5) | 1.2 (0.9, 1.5) | 0.29 | 0.1362 |
| Gastrointestinal or liver disease | 140 (4.7) | 52 (7.8) | 1.5 (1.2, 2) | 1.4 (1, 2) | **0.04** | 0.1736 |

Abbreviations: COVID-19 = coronavirus disease 2019; COVID-NET = COVID-19–Associated Hospitalization Surveillance Network; LTCF = long-term care facility
<sup>a</sup>Log-linked Poisson Regression using generalized estimating equations (GEE) clustered on site with exchangeable covariance structure
<sup>b</sup>California, Connecticut, Colorado, Georgia, Maryland, Michigan, Minnesota, New Mexico, New York, Ohio, Oregon, Tennessee, and Utah.
<sup>c</sup>Race and Hispanic ethnicity: Data on race and ethnicity were categorized as follows: non-Hispanic White (White), non-Hispanic Black (Black), Other/Unknown. If ethnicity was unknown (4%), non-Hispanic ethnicity was assumed.
<sup>d</sup>Includes American Indian/Alaska Native, Asian/Pacific Islander, and multiple/unknown race.
<sup>e</sup>Long-term care facility (LTCF) residence was defined as residence in rehabilitation facilities, assisted living/residential care, group homes, nursing homes, skilled nursing facilities, long-term care facilities, long-term acute care hospitals, residential care facilities or other long-term care facilities.
<sup>f</sup>Cardiovascular disease excludes hypertension.

**Supplementary Table 3B.** Multivariable model^a^ assessing factors associated with severe COVID-19 diseases (ICU, in-hospital death) in hospitalized adults with laboratory-confirmed COVID-19-associated hospitalization admitted January 1 – June 30, 2021, restricting to those with COVID-19-associated hospitalization as the likely primary reason for admission, among those aged ≥65 years — COVID-NET, 13 States^b^.

| Category | No severe illness<br>n (weighted %) | Have severe illness<br>n (weighted %) | Unadjusted risk ratio | Adjusted risk ratio | p-value | Standard error |
| --- | --- | --- | --- | --- | --- | --- |
| **Race and Hispanic ethnicity<sup>c</sup>** |  |  |  |  |  |  |
| White | 829 (60.5) | 231 (50.0) | R | R | R | R |
| Black | 185 (21.5) | 77 (28.4) | 1.4 (0.9, 2.3) | 1.6 (1, 2.6) | **0.05** | 0.2433 |
| Hispanic or Latino | 89 (7.4) | 45 (10.1) | 1.4 (1, 2.1) | 1.6 (1.2, 2.3) | **0.005** | 0.1736 |
| Other/Unknown<sup>d</sup> | 101 (10.6) | 44 (11.5) | 1.2 (0.9, 1.7) | 1.3 (0.9, 1.8) | 0.15 | 0.1722 |
| **Sex** |  |  |  |  |  |  |
| Male | 576 (52.5) | 228 (55.9) | 1.1 (0.8, 1.5) | 1.1 (0.8, 1.4) | 0.47 | 0.1326 |
| Female | 628 (47.5) | 169 (44.1) | R | R | R | R |
| **Age (in years)** |  |  |  | 1 (1, 1) | 0.34 | 0.0086 |
| **Unvaccinated** | 1,051 (94.8) | 350 (94.6) | R | R | R | R |
| **Fully vaccinated** | 153 (5.2) | 47 (5.4) |  | 1.1 (0.7, 1.6) | 0.80 | 0.2167 |
| **LTCF residence<sup>e</sup>** | 90 (10.3) | 44 (14.2) | 1.3 (0.9, 1.9) | 1.5 (1, 2.2) | 0.06 | 0.2019 |
| **Underlying medical condition** |  |  |  |  |  |  |
| Neurologic disease | 304 (29.0) | 94 (22.5) | 0.8 (0.6, 1) | 0.7 (0.5, 0.9) | **0.02** | 0.1486 |
| Immunosuppressive condition | 155 (13.0) | 59 (18.5) | 1.4 (1.1, 1.7) | 1.4 (1.1, 1.7) | **0.002** | 0.1097 |

Abbreviations: COVID-19 = coronavirus disease 2019; COVID-NET = COVID-19–Associated Hospitalization Surveillance Network; LTCF = long-term care facility
<sup>a</sup>Log-linked Poisson Regression using generalized estimating equations (GEE) clustered on site with exchangeable covariance structure
<sup>b</sup>California, Connecticut, Colorado, Georgia, Maryland, Michigan, Minnesota, New Mexico, New York, Ohio, Oregon, Tennessee, and Utah.
<sup>c</sup>Race and Hispanic ethnicity: Data on race and ethnicity were categorized as follows: non-Hispanic White (White), non-Hispanic Black (Black), Other/Unknown. If ethnicity was unknown (7% of cases), non-Hispanic ethnicity was assumed.
<sup>d</sup>Includes American Indian/Alaska Native, Asian/Pacific Islander, and multiple/unknown race
<sup>e</sup>Long-term care facility (LTCF) residence was defined as residence in rehabilitation facilities, assisted living/residential care, group homes, nursing homes, skilled nursing.

**Supplementary Table 4.** Rate ratios of COVID-19-associated hospitalizations in unvaccinated vs vaccinated patients among all adults aged ≥18 years and by age group ^a^, January 24–July 24, 2021^b^ – COVID-NET, 13 States^c^.

| | Rate ratio of hospitalizations<br>all adults $\geq 18$ years (age-adjusted) | Rate ratio,<br>18–49 years | Rate ratio,<br>50–64 years | Rate ratio,<br>$\geq 65$ years |
| --- | --- | --- | --- | --- |
| **MMWR week ending** |  |  |  |  |
| 1/30/2021 | 21 | - | 7 | - |
| 2/6/2021 | 13 | - | - | 7 |
| 2/13/2021 | 8 | 10 | 26 | 5 |
| 2/20/2021 | 8 | 23 | 21 | 4 |
| 2/27/2021 | 6 | 15 | 23 | 4 |
| 3/6/2021 | 14 | 21 | 15 | 11 |
| 3/13/2021 | 10 | 52 | 14 | 6 |
| 3/20/2021 | 12 | 15 | 12 | 11 |
| 3/27/2021 | 20 | 21 | 22 | 18 |
| 4/3/2021 | 15 | 75 | 15 | 11 |
| 4/10/2021 | 17 | 45 | 22 | 13 |
| 4/17/2021 | 28 | 18 | 53 | 25 |
| 4/24/2021 | 24 | 40 | 51 | 16 |
| 5/1/2021 | 22 | 33 | 31 | 17 |
| 5/8/2021 | 31 | 51 | 33 | 26 |
| 5/15/2021 | 26 | 32 | 49 | 18 |
| 5/22/2021 | 19 | 13 | 23 | 21 |
| 5/29/2021 | 21 | 19 | 16 | 25 |
| 6/5/2021 | 29 | 24 | 78 | 23 |
| 6/12/2021 | 15 | 17 | 21 | 13 |
| 6/19/2021 | 17 | 13 | 25 | 16 |
| 6/26/2021 | 17 | 12 | 16 | 22 |
| 7/3/2021 | 14 | 13 | 18 | 12 |
| 7/10/2021 | 12 | 19 | 13 | 10 |
| 7/17/2021 | 17 | 17 | 20 | 15 |
| 7/24/2021 | 19 | 20 | 25 | 16 |
| **Cumulative** | **17** | **23** | **22** | **13** |
| **Range** | **6–31** | **10–75** | **7–78** | **4–26** |

Abbreviations: COVID-19 = coronavirus disease 2019; COVID-NET = COVID-19–Associated Hospitalization Surveillance Network
<sup>a</sup>Age-specific rate ratios could not be calculated for weeks during which zero vaccinated cases were reported and are represented by a dash (-).
<sup>b</sup>COVID-NET hospitalization data are preliminary and subject to change as more data become available. In particular, rates for recent hospital admissions are subject to lag.
<sup>c</sup>California, Connecticut, Colorado, Georgia, Maryland, Michigan, Minnesota, New Mexico, New York, Ohio, Oregon, Tennessee, and Utah.

### Supplementary Figures

**Supplementary Figure 1.**
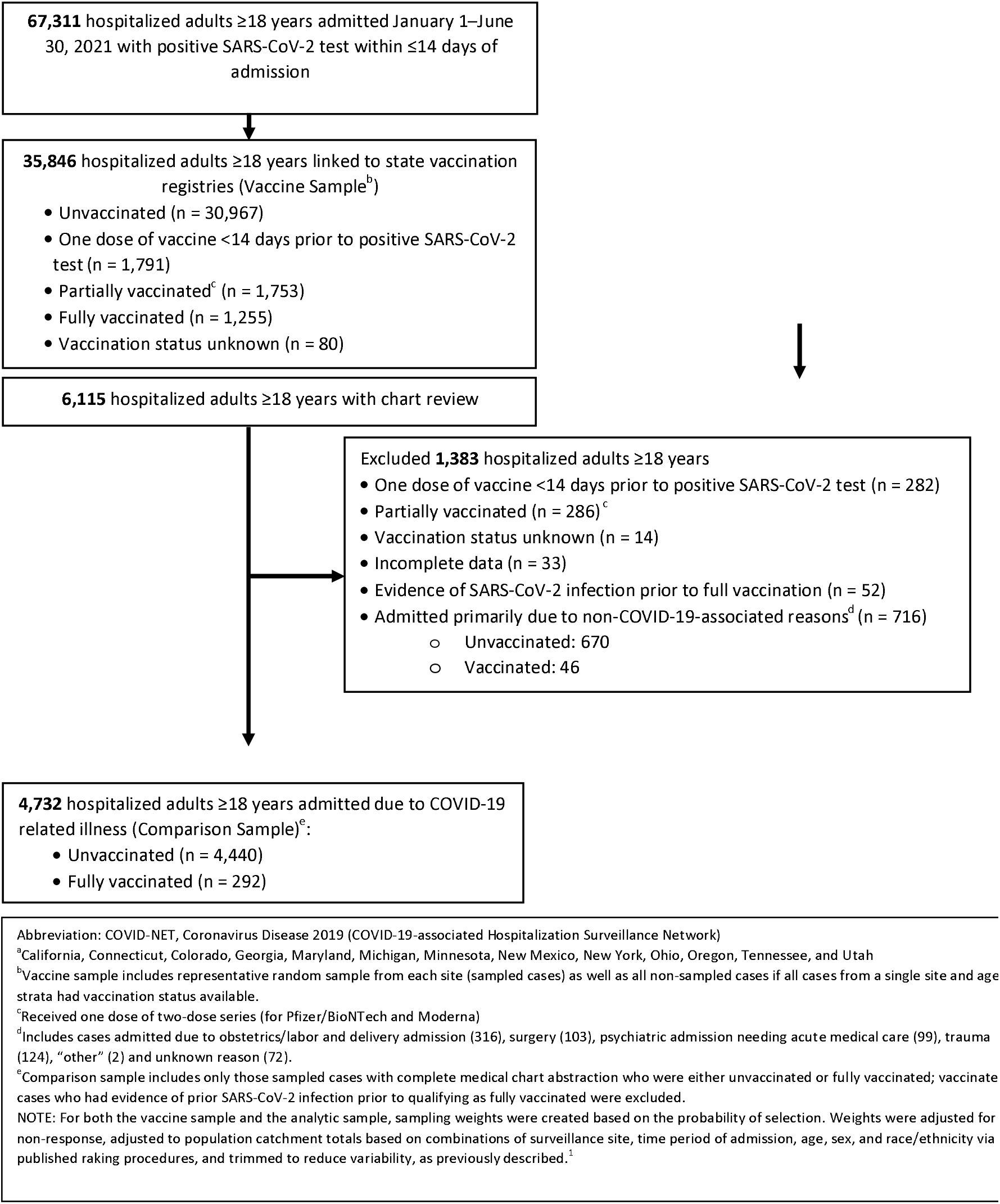
Selection of cases for analysis of adults ≥18 years with laboratory-confirmed COVID-19-associated hospitalization admitted January 1 – June 30, 2021, by vaccination status — COVID-NET, 13 States^a^.

**Supplementary Figure 2.**
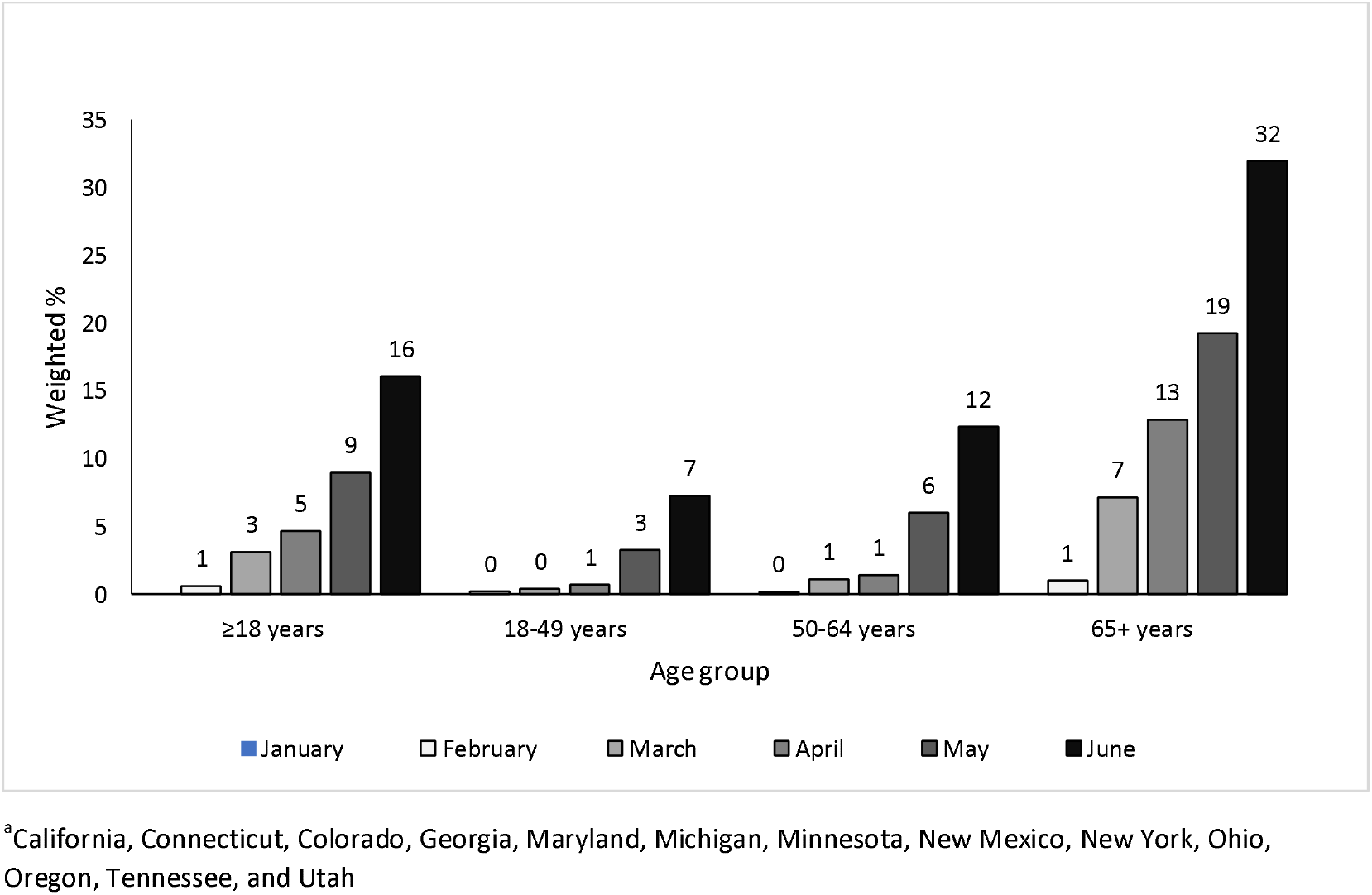
Proportion of adults ≥18 years with COVID-19-associated hospitalizations admitted January 1 – June 30, 2021 who are fully vaccinated, by age group and month of admission – COVID-NET, 13 States^a^.

**Supplementary Figure 3.**
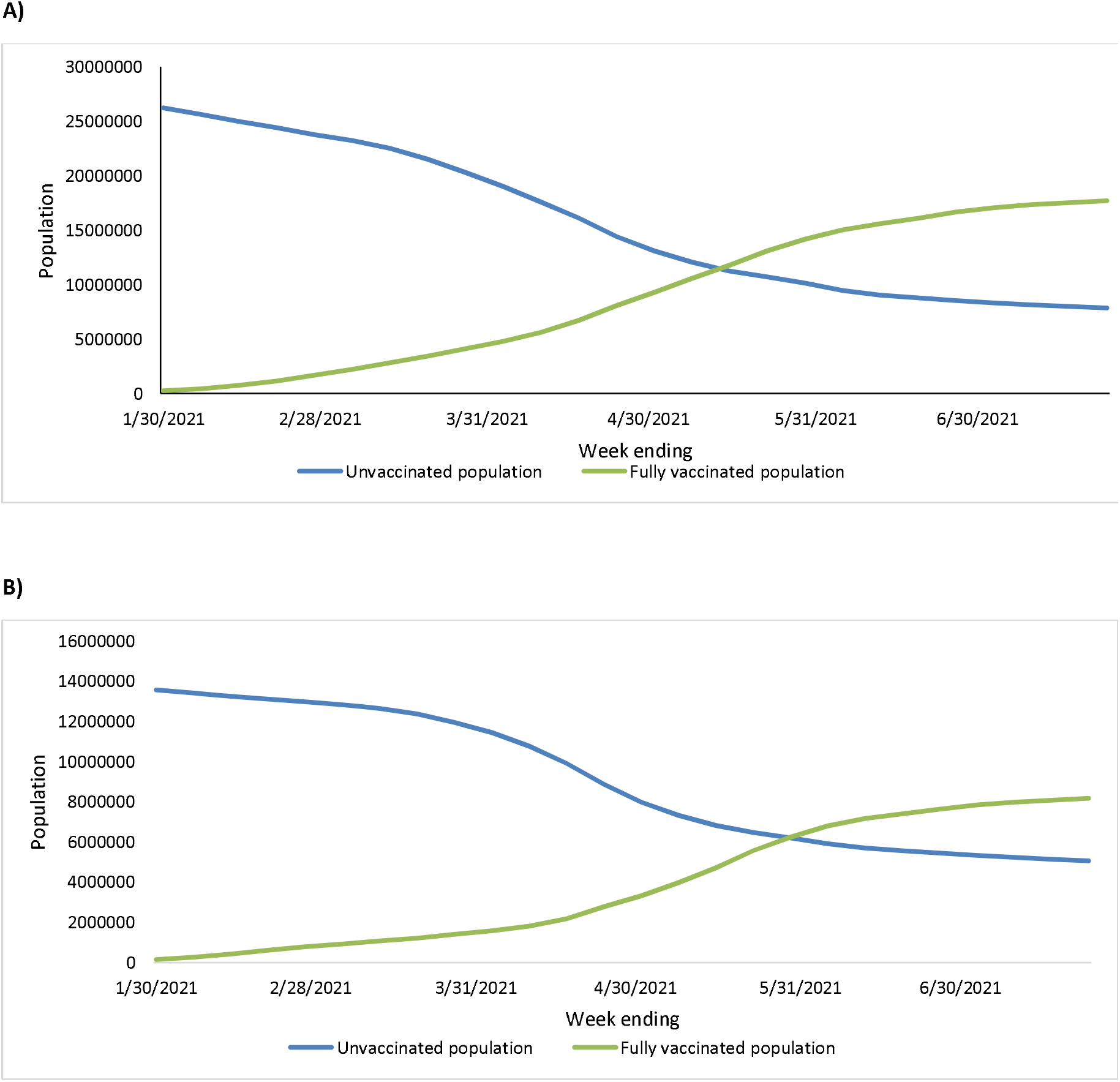
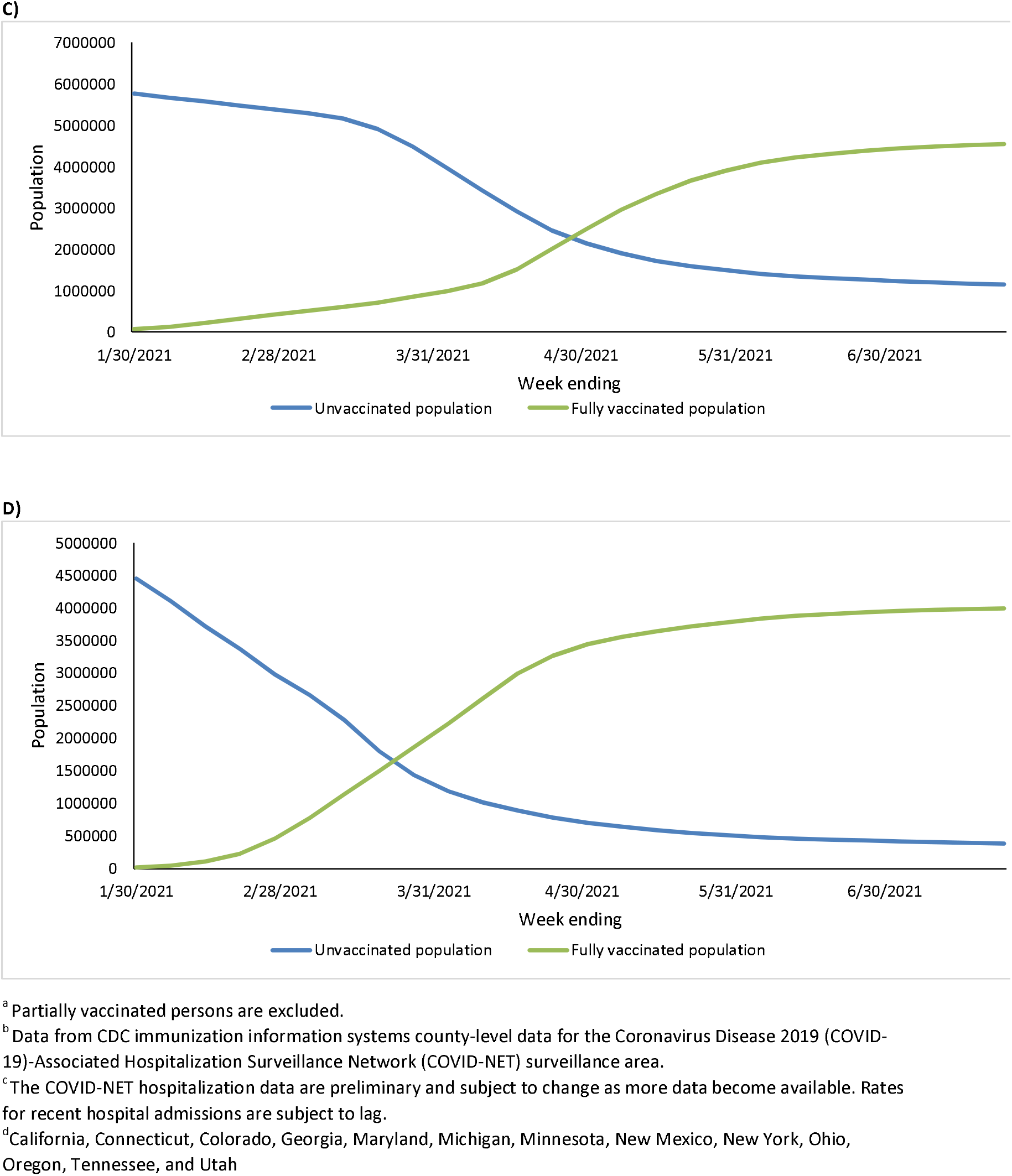
COVID-NET catchment population by vaccination status^a^ and week, used as denominator in population-based-rate calculations^b^ among A) all adults ≥18 years, B) 18–49 years, C) 50–64 years, and D) ≥65 years, January 24–July 24, 2021—COVID-NET^c^, 13 States^d^.

